# Deep brain stimulation surgery under ketamine induced conscious sedation: a double blind randomized controlled trial

**DOI:** 10.1101/2023.08.26.23294660

**Authors:** Evgeniya Kornilov, Halen Baker Erdman, Eilat Kahana, Shlomo Fireman, Omer Zarchi, Michal Israelashvili, Johnathan Reiner, Amir Glik, Penina Weiss, Rony Paz, Hagai Bergman, Idit Tamir

## Abstract

**Background:** The gold standard anesthesia for deep brain stimulation (DBS) surgery is the “awake” approach, using local anesthesia alone. While it offers high-quality microelectrode recordings and clinical assessment of the stimulation therapeutic window, it potentially causes patients extreme stress and might result in suboptimal surgical outcomes. However, the alternative of general anesthesia or deep sedation dramatically reduces reliability of physiological navigation and therapeutic window assessment, thus potentially diminishing the accuracy of lead localization. We therefore designed a prospective double-blinded randomized controlled trial to investigate a novel anesthesia regimen of ketamine-induced conscious sedation for DBS surgery.

**Methods:** Patients with Parkinson’s disease undergoing subthalamic nucleus DBS surgery were enrolled. Patients were randomly assigned to either the experimental or control group. During the physiological navigation phase, the experimental group received ketamine infusion at a dosage of 0.25 mg/kg/hr, while the control group received normal saline. Both groups received moderate propofol sedation before and after the physiological navigation phase. Primary outcomes were non-inferiority of electrophysiological quality, including multiunit recordings, EEG, EMG, bispectral index and lead localization accuracy according to postoperative CT scans. Secondary outcomes included patients’ satisfaction level measured using Iowa satisfaction with anesthesia scale for awake procedures. Potential side effects and adverse events were also monitored, including hemodynamics (blood pressure, heart rate) and cognition (hallucinations during surgery and early post-operative cognition using Montreal Cognitive Assessment).

**Results:** Thirty patients, 15 from each group, were included in the study and analysed. Intra-operatively, the electrophysiological signature of the subthalamic nucleus was similar under ketamine and saline. Tremor amplitude was slightly lower under ketamine (*p* = 0.002). The accuracy of lead position was comparable in both groups. Postoperatively, patients in the ketamine group reported significantly higher satisfaction with anesthesia. The improvement in Unified Parkinson’s disease rating scale part-III was similar between the groups. No negative effects of ketamine on hemodynamic stability or cognition were reported perioperatively. Additionally, no procedure-related complications were reported in either group, besides one case of peri-lead edema in the control group.

**Conclusion:** This study demonstrates that ketamine induced conscious sedation during physiological navigation in DBS surgery resulted in non-inferior intra-operative, post-surgical and patient satisfaction outcomes compared to the commonly used standard awake protocol, without major disadvantages. Future studies should investigate the applicability of this protocol in other awake neurosurgical procedures, such as DBS for other targets and indications, and awake craniotomy for tumor resection and epilepsy.

## Introduction

Deep Brain Stimulation (DBS) surgery has been performed ‘awake’ (under local anesthesia only) for many years. The purpose of having the patient awake was to enable microelectrode recordings (MER) to verify target localization, and to clinically assess the therapeutic window (TW) by intra-operative stimulation at the target. Commonly used sedatives and anesthetic agents significantly alter the recorded brain signal, the patient’s symptoms, and the ability of the patient to cooperate with test stimulation.^1–4^ Thus, they are less suitable for this surgery.

Awake surgery is a major obstacle of DBS, both for the patient and the operating room staff. It is fairly common for awake DBS cases to be altered or aborted, due to patient’s extreme anxiety and inability to cooperate. This may results in sub-optimal surgical outcome due to lead location inaccuracy, unfavorable clinical outcome, traumatic patient experience,^5^ or other surgical complications like intracranial bleeding.

As technology develops over the years, higher resolution and improved signal-to-noise ratio MRI and CT scans are now available, including scanners for intra-operative guidance. Additionally, innovative neuronavigational systems allow for better fusion and registration protocols. Therefore, the accuracy of lead implantation has increased over the years, and the need for ‘awake’ surgery is now questioned. Finally, segmented leads allow more flexibly in current steering that can compensate for less optimal lead location. A growing number of centers have changed their practice to general anesthesia, preferring patient and staff comfort over MER and clinical assessment. While DBS under general anesthesia is doable, the cost-benefit for a single patient undergoing elective surgery should be carefully considered. The borders of the DBS target (e.g., the motor subdomain of the subthalamic nucleus) still cannot be clearly deduced from today’s standard clinical imaging.^6,7^ In addition, other factors can contribute to electrode misplacement, among these are stereotactic errors, brain shift and more.^8–13^ Intra-operative MER and clinical assessment can help overcome these issues by enabling real-time identification and correction of misplaced DBS leads.

We recently published a novel protocol for a well-known drug – ketamine, to be used at a low dose to create conscious dissociative sedation and enable MER and TW assessment.^14,15^ The first study, conducted in non-human primates, showed the ability to record spiking activity in the basal ganglia and the frontal cortex under ketamine sedation, with similar quality to ‘awake’ recordings. We then used a comparable ketamine-based anesthesia protocol in humans and retrospectively showed the ability to record spiking activity under ketamine in patients undergoing DBS surgery. The protocol involved administration of propofol for the initial phase of the surgery (skin incision and burr hole drilling) and ketamine for the subsequent phase (MER and clinical assessment). Finally, propofol is given in the third phase of surgery (lead fixation and skin closure). There were no adverse events, nor complications attributed to ketamine use. The overall experience of the patients under ketamine seemed positive, likely due to its analgetic and anxiolytic properties. However, due to the retrospective nature of the study, there was no formal assessment of the patients’ experience and no unity in the ketamine dose used. We therefore designed a prospective single-center double blind randomized controlled trial to assess the non-inferiority of ‘propofol-ketamine’ over ‘propofol-awake’ anesthesia regimen.

## Materials and methods

### Study design

This study was conducted at Beilinson Campus of the Rabin Medical Center, Petah Tikva, Israel between September 2020 and January 2022 and was designed as a single center prospective, double-blind randomized placebo-controlled interventional trial. The study protocol was approved by the Institutional Ethics Committee and registered at https://clinicaltrials.gov (NCT04716296). All participants obtained and provided written informed consent.

### Patient selection and inclusion criteria

Patients with Parkinson disease (PD), having movement disorder society - Unified Parkinson’s disease Rating Scale (MDS-UPDRS) part III improvement of >30% in levodopa-challenge test, and otherwise eligible and planned for DBS surgery targeting the subthalamic nucleus (STN), participated in the study. If any adverse event happened during surgery including patient’s extreme discomfort or anxiety – the anesthesia regimen was changed accordingly, and the patient was excluded from data analysis.

### Outcomes

#### Primary outcomes

1. Non-inferiority of the electrophysiological properties of MER in the STN during ketamine administration compared to a placebo.
2. Comparison of EEG, EMG and bispectral index (BIS) during the surgery in two study groups.

#### Secondary outcomes

1. Evaluation of side effects during ketamine administration (hypertension, tachycardia, hemodynamic instability, hallucinations).
2. Evaluation of early post-operative cognitive decline by Montreal Cognitive Assessment (MoCA) scale on the second postoperative day.
3. Patients’ satisfaction level measured using ISAS (Iowa satisfaction with anesthesia scale) for awake procedures.

### Sample size calculation

Sample size calculation was based on STN length identified with electrophysiology during DBS surgery. It usually fluctuated between 4 and 6 mm with a standard deviation (SD) of 1.6 mm.^16,17^ The minimal STN length used for favorable DBS outcome is 4 mm.^17^ Using significance level of 5% (alpha) and power of 80% with SD of 1.6 mm and non-inferiority limit of 1.5 mm, a total 30 patients were required to prove no difference between placebo (saline) and experimental (ketamine) anesthesia protocols, 15 patients per group. Taking into account the follow-up period of 3 to 6 months, 15% of potential dropouts were allowed, and 35 patients were included in the study.

### Randomization

Using block randomization technique, 17 patients were allocated to the experimental group and 18 to the control group. A computer-generated randomization list was prepared by a research assistant in opaque, sealed and numbered envelopes containing the group assignment. After inclusion, patients were given a number in the order of their participation in the study. On the morning of surgery, the neurosurgeon opened the corresponding numbered envelope to determine the patient’s group assignment (Group1 or Group2) and informed the anesthesiologist of the appropriate anesthesia protocol.

### Blinding

Blinding of all the involved treatment providers (including the neurosurgeon and anesthesiologist) was not ethically feasible in our study. The neurosurgeon and the anesthesiologist were therefore not blinded. Patients, neurophysiologists, and the movement disorder neurologist were blinded throughout the duration of the study. Data analysis was made on blinded data, with the groups only being revealed upon completion of the analysis.

### Surgery

DBS surgery targeting the STN, was performed as previously described.^15^ Briefly, the STN was identified with high resolution 3T T2-weighted preoperative MRI. Trajectory to target was planned using T1 with gadolinium, 3D volumetric 1 mm slices protocol. Parkinson’s medications were discontinued the evening before surgery. On the morning of surgery, a stereotactic frame (CRW, Integra) was fixated on the patient’s head under local anesthesia (lidocaine 2% and bupivacaine 0.5%, ∼20 cc) and mild sedation (IV midazolam 2 mg). A CT scan was then performed with a fiducial box attached to the stereotactic frame, and the CT and MRI images were fused using Medtronic Stealth 8 navigation system to extract precise coordinates for electrode implantation. The patient was transferred to the operating room (OR), positioned with his head elevated 15 to 30 degrees, and the frame was securely attached to the operating table. An intra-operative CT (O-Arm, Medtronic) was brought into the operating theatre and was located around the patient’s head throughout the procedure.

The surgical procedure consisted of three stages:

1. After skin incision, a burr hole was made in the skull at the desired entry point, then the lead fixation device was mounted to the skull and a brain cannula was lowered to a depth of 15 mm above the target.
2. Recording microelectrodes (Sonus, Alpha Omega Engineering, Ziporit, Israel, typical impedance at 1000Hz = 0.3-0.7 MΩ) were lowered through the cannula to 10 mm above target, and MER was performed by a neurophysiologist, aided by a Hidden Markov Model (HMM) algorithm for STN detection (HaGuide, Alpha Omega Engineering, Ziporit, Israel).^18,19^ Monopolar cathodal stimulation (130 Hz, 60 μS pulse width, 0.25-4 mA) through a macro-contact (1.25 mm diameter, 1 mm length, typical impedance at 30Hz =1-3 kΩ) located 3 mm above the micro-contact, was applied at the lower border of the dorso-lateral oscillatory region (DLOR) of the STN to evaluate the TW.
3. If MER recording reveled >4 mm STN length and the TW was found to be acceptable, the recording electrodes were replaced by a permanent DBS lead (3389, Medtronic Inc.), usually with the bottom of the second contact from below at the stimulation depth, i.e., lower border of the DLOR. The lead was fixated to the skull-mounted lead fixation device (Stimloc, Medtronic) and intra operative CT (O-arm, Medtronic) confirmed lead location. Upon confirmation, the skin was closed.

The procedure was carried out for one or both hemispheres. Moderate sedation with propofol was resumed for the lead fixation and skin closure. Directly after, the stereotactic frame was removed from the patient’s head, the patients underwent intubation and general anesthesia and the lead extenders were connected to the brain leads and the internal pulse generator was connected and implanted on the patient’s chest.

### Anesthesia protocols for experimental and control groups

Upon entering the OR, patients were given IV paracetamol (1 g) and ondansetron (4 mg), an 18-gauge IV line was established in addition to the previously inserted 20-gauge IV line and cannulation of radial artery (20-gauge catheter) was performed for blood pressure management. All patients were monitored in accordance with The American Society of Anesthesiologists (ASA) guidelines (ECG, Sp02, non-invasive and invasive blood pressure monitoring, and nasal EtCO_2_ monitoring (iMDsoft’s MetaVision® and Mindray^©^)). Additionally, a BIS monitor (BIS Vista, Covidien Medtronic Dublin, Ireland) was utilized. Antibiotic prophylaxis was administered 30 minutes prior to surgery (either cefazoline 2 g or clindamycin 900 mg).

Initial (before sedation) BIS values were between 92-96. Throughout the first and third stages of surgery, all patients received a continuous propofol infusion to maintain moderate sedation (BIS 60-80). Fifteen minutes prior to the second stage of surgery (MER and clinical assessment), when the burr hole was ready, the propofol infusion was discontinued, and patients received either a placebo (normal saline) or an interventional drug (ketamine), depending on their assigned group:

- The ketamine group received a continuous 0.1% ketamine infusion, (Renaudin, France), at a dosage of 0.25 mg/kg/hr (rate of 0.25ml/kg/hr).
- The saline group received a continuous normal saline (NaCl 0.9%) infusion at a rate of 0.25 ml/kg/hr.

Upon completion of the second stage of surgery, the placebo/interventional drug was discontinued and propofol was resumed until the end of the third stage. Intraoperative blood pressure was closely monitored and maintained within accepted limits (mean arterial pressure 65-85 mmHg, systolic blood pressure < 150 mmHg). In cases of hypertension, treatment was administered in the form of nicardipine, hydralazine or labetalol.

Pulse generator implantation was performed using a standard general anesthesia technique. Anesthesia induction consisted of propofol (1-2 mg/kg), fentanyl (0.1-0.2 mg) and rocuronium (30-50 mg) followed by maintenance with sevoflurane (minimum alveolar concertation 1-1.2). After surgery completion, patients were extubated, transferred to the post-anesthesia care unit (PACU) and transferred to the neurosurgical department the following day. PACU management included administration of IV fluids, antibiotics, analgetic and antiemetic drugs as needed, along with anti-parkinsonian treatment.

### Follow-up

All patients underwent a preoperative evaluation (2 months before the surgery) and were subsequently monitored 3-6 months after surgery. The post-op evaluation included UPDRS part III and L-dopa equivalent daily dose (LEDD). Cognitive impairment screened with MoCA test was assessed preoperatively (1-2 months before the surgery) as well as on the second postoperative day to measure early cognitive decline. To gain a comprehensive understanding of patients’ experience during the “awake” (experimental) part of their surgery, we used the ISAS (Iowa Satisfaction Anesthesia Scale),^20^ a validated questionnaire that measures patient satisfaction with monitored anesthesia care during awake surgeries. Patients were asked to complete the questionnaire within 24-48 hours postoperatively and to refer only to their experience of the experimental part of surgery.

### Electrophysiological data collection

Data acquisition for neurophysiological signals (MER, EEG and EMG), was performed utilizing the Neuro-Omega navigation system (Alpha Omega Engineering. Ziporit, Israel), offering precise spatial control of the micro-electrode position and real-time monitoring.

Multi-unit recordings were obtained through the microelectrodes (average impedance at 1000Hz = 0.5 ± 0.12 MΩ), which were inserted into the target area. Recording began 10 mm above target, advancing in increments of 0.4 mm before STN and 0.1 mm within the nucleus. Each site was recorded for at least 4 seconds after 2 seconds of stabilization. EEG electrodes were located at Fp1, Fp2, TP9 and TP10 locations according to the 10-20 system. EMG sensors were attached to the patient’s forearm to collect signals from the flexors (flexor carpi radialis) and extensors (extensor carpi radialis) muscles on both the right and left arms.

### Data analysis

All intraoperative data (except spiking data that were recorded only in the experimental stage), was divided into 4 time periods: baseline (patient awake in the OR before propofol infusion), propofol infusion during the first surgery stage (propofol-1); experimental stage during MER (ketamine/saline), and propofol administration during third surgery stage during lead fixation and scalp closure (propofol-2). See Supplementary Fig. 1 for the details.

Spiking data (sampled at 44kHz, 300-6000 Hz SW band-pass filtered, following HW 0.1-9000Hz band-pass filter), recorded from the microelectrode trajectory, was extracted from the Neuro-Omega system. The borders and subdomains of the STN were determined using a previously published algorithm.^19,21^ Briefly, the power spectrum density (PSD) and RMS were calculated for each recording site. The RMS was then normalized by the first 6 pre-STN (internal capsule white matter) sites (baseline RMS), resulting in the normalized RMS (NRMS). The peak value (peak RMS) and area under the curve (RMS AUC) of NRMS were computed for each trajectory. Spectrograms of the discharge rate (following absolute operator to expose low-frequency oscillations) of individual patients were normalized by the total power per site and calculated. The length of the STN was extracted and the NRMS and the spectrogram were normalized by the patient’s STN length and then grouped and averaged for each patient group. Percentage STN designated DLOR (DLOR %) by the HMM algorithm and confirmed by an expert electrophysiologist was calculated as: DLOR length/STN length*100. The ratio of beta power (intensity of beta oscillations) within the STN to beta power outside the STN was calculated for each trajectory.

EEG data was sampled at 1000Hz, software band-passed between 1 and 150 Hz with 4^th^ order Butterworth filter. 50 Hz notch filter was applied to remove electrical noise. The difference between frontal and temporo-parietal electrodes (Fp1 - TP9 and Fp2 - TP10) was taken from each side. Power spectral density was calculated using Welch’s method with Hamming window of 3 seconds (yields a resolution of 1/3 Hz), and 2/3 overlap. EEG spectra were compared using 75% spectral edge frequency (SEF75) and also with area under the curve (AUC) for each EEG frequency band including delta (1-4 Hz), theta (4-8 Hz), alpha (8-13 Hz), beta (13-30 Hz), gamma (30-70 Hz), high frequency oscillations (HFO, 70-150 Hz).

EMG data with initial sampling rate of 11 kHz was resampled to 1.1 kHz and band-passed between 10 and 200 Hz with 4^th^ order Butterworth filter and rectified. PSD was calculated for each EMG electrode using Welch’s method with Hamming window of 3 seconds, and 2/3 overlap (1/3 Hz resolution). The maximal power of the frequency band typical for parkinsonian tremor (4-6 Hz) was compared between the groups.

BIS measurements were extracted from the BIS device and resampled to 1/60 Hz (1 measure per minute). BIS values from separate patients were linearly interpolated to the same time length for each time period. Averaged BIS values for each of the periods were compared between experimental and control groups.

Hemodynamics measurements consisted of systolic blood pressure (SBP), mean arterial pressure (MAP) and heart rate (HR). Blood pressure values measured by arterial-line and heart rate – by ECG, were extracted from MetaVision® system and underwent similar preprocessing as for BIS values.

Because of possible sympathomimetic properties of ketamine,^22^ it was decided to measure hemodynamics variability and hypertensive/tachycardic events in the study cohort. Variability was assessed with two common methods:^23,24^ coefficient of variation (CV; standard deviation divided by the corresponding mean) and average real variability (ARV; average of the absolute differences between all consecutive BP/HR measurements). Hypertensive/tachycardic event was defined as an increase in BP/HR more than 2 standard deviations of averaged BP/HR in one minute. The frequency of these events was calculated for every patient (number of events divided by length of each time period in minutes). All coefficients were calculated separately for each time period as well as for the whole experiment for each patient.

ISAS questionnaires were analyzed in accordance with the instructions (mean response of all 11 questions), with maximal score of 3 (maximal satisfaction) and minimal of −3.

The accuracy of lead implantation at the target was evaluated by measuring the radial error. The radial error was defined as the distance between the center of the implanted lead at the target plane and the planned target point. This was measured in probe’s eye view at 90° to the trajectory plane on the navigation system, using the fused images of the intra-operative CT and pre-operative MRI.

All analysis was performed using custom Matlab (R2017a or R2021a) scripts.

### Statistical analysis

After data preparation, the statistical assumptions were examined. For data that followed a normal distribution, parametric tests were used, while non-parametric tests were employed for non-normally distributed data. The results were presented as mean and standard error of mean (SEM) for normally distributed continuous data, median (interquartile range) for non-normally distributed data, and count (percentage) for categorical variables. To compare groups, the Welch test or Mann-Whitney U test was used for continuous data, and the Chi-squared test for categorical data. Significance was determined as a p-value of less than 0.05 (two-tailed) except for ISAS, where right-tailed Mann-Whitney test was used (since ketamine has anxiolytic and anti-depressive properties) to test for more anesthesia satisfaction in ketamine group. Bonferroni correction was used in case of multiple comparisons (p-value of 0.05 divided by number of comparisons).

### Data availability

The data that support the findings of this study are available from the corresponding author, upon reasonable request.

## Results

A total of 35 patients participated in the study. After exclusion, 30 patients, 15 from each group, were included in the analysis (Fig. 1). Brief experimental design with typical examples of data collection is depicted in Supplementary Fig. 1. Perioperative patient characteristics (demographic and disease characteristics) showed no difference between the groups (Table 1).

**Figure 1.**
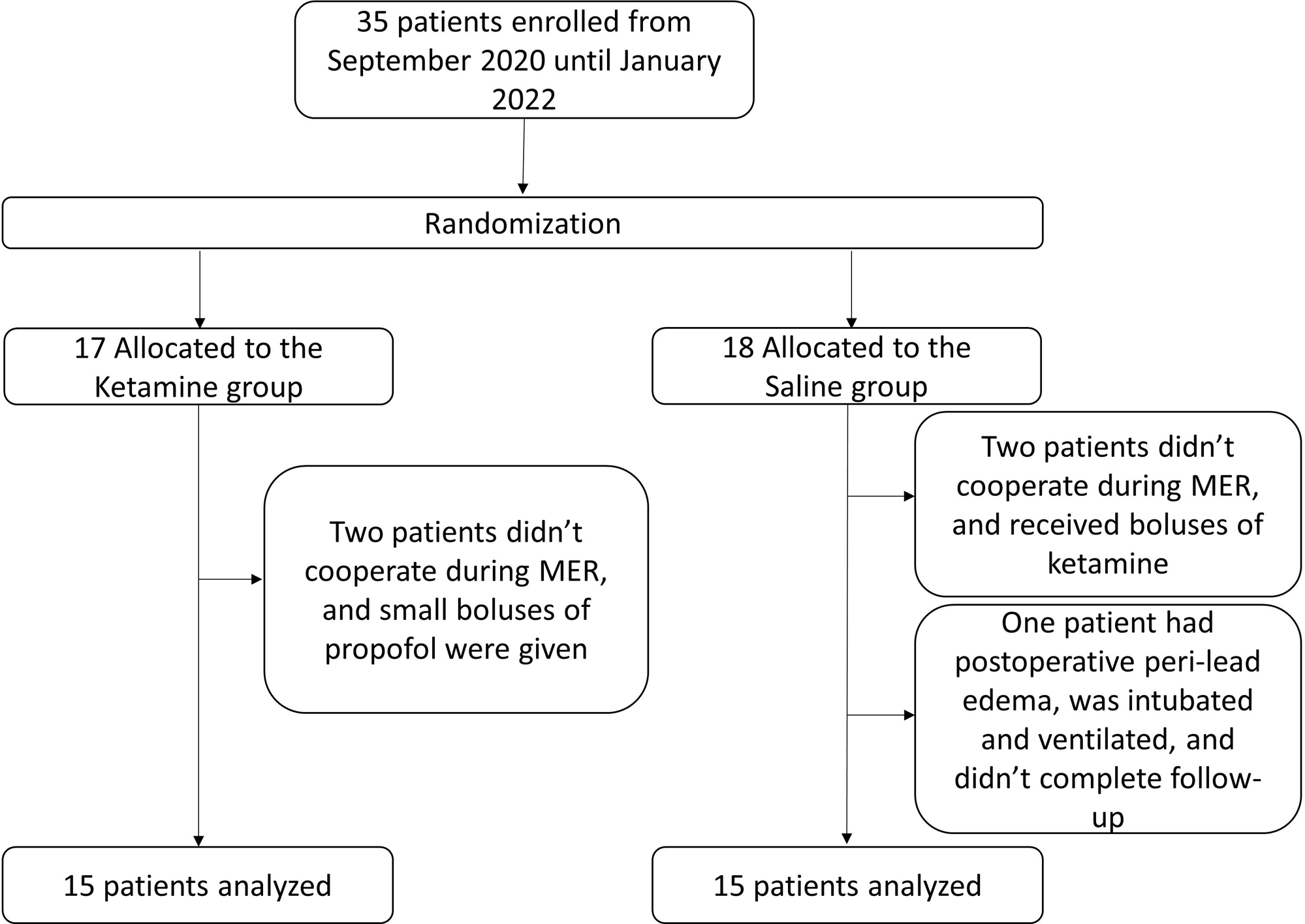
Patient flowchart. MER – microelectrode recordings.

**Table 1.** Demographic data, pre-operative, intra-operative and post-operative patient characteristics. Data presented as median (interquartile range). P-values for continuous variables are calculated with Mann-Whitney test; and for categorical variables with chi-square test. a - Ketamine was given at a fixed rate – 0.25 mg/kg/hr. LEDD - levodopa equivalent daily dose; MMSE - Mini-Mental State Examination; MoCA - Montreal Cognitive Assessment; UPDRS-III Unified Parkinson’s disease rating scale, Part 3.

|  | Ketamine group, N = 15 | Saline group, N = 15 | P-value |
| --- | --- | --- | --- |
| **Patients' demographics** |  |  |  |
| Age (years) | 67 (57.75-74) | 69 (64.25-74.5) | 0.28 |
| Weight (kg) | 70 (64-72) | 73 (64.25-84.75) | 0.589 |
| Male sex | 10(66.7%) | 10(66.7%) | 1 |
| Disease duration (years) | 7 (4.25-8) | 6.5 (5-10) | 0.455 |
| **Preoperative data** |  |  |  |
| LEDD pre (mg) | 1100 (550-1555) | 925 (500-1406) | 0.589 |
| UPDRS-III preoperative, off medication | 46.5 (42-55.5) | 43 (24.5-50) | 0.231 |
| UPDRS-III preoperative, on medication | 24.5 (15-30.5) | 19 (8-30) | 0.395 |
| MoCA 2 months preoperative | 24 (19-27) | 25 (23-28) | 0.32 |
| MMSE preoperative | 28 (24.25-29) | 29 (28-30) | 0.051 |
| Beck's Anxiety Inventory, preoperative | 16 (7.75-18.5) | 10.5 (6-17) | 0.641 |
| Beck's Depression Inventory, preoperative | 12 (8.75-21.25) | 10.5 (7-15) | 0.609 |
| **Intraoperative data** |  |  |  |
| Propofol drip (mg/kg/hr) | 3.7 (3.4-4.7) | 3.4 (2.5-4.9) | 0.361 |
| Propofol total (mg) | 691 (509.75-938.5) | 550 (452.75-778.75) | 0.245 |
| Ketamine (mg)<sup>a</sup> | 44 (30.25-55.75) | NR | NR |
| Nicardipine –total (mg) | 8.1 (2.825-12.475) | 5.7 (4.325-13.45) | 0.983 |
| Labetalol – total (mg) | 0 (0-10) | 0 (0-3.75) | 0.29 |
| Hydralazine – total (mg) | 4 (0-11.875) | 0 (0-7.5) | 0.26 |
| Anaesthesia duration (minutes) | 329 (285.5-361.25) | 294 (274.25-354.25) | 0.575 |
| Surgery duration (minutes) | 244 (215.5-268.75) | 243 (210.5-287) | 1 |
| **Postoperative data** |  |  |  |
| Radial error (mm) | 0.8 (0.375-1) | 0.6 (0.325-0.9) | 0.59 |
| MoCA, second postoperative day | 22 (18.25-25.75) | 23.5 (21-25) | 0.463 |
| UPDRS-III postoperative on medication, on stimulation | 9 (6-19) | 11 (6-14) | 0.856 |
| LEDD post (mg) | 450 (145.75-627) | 440 (206.25-594) | 0.764 |
| Length of hospital stay, days | 3 (3-4) | 3 (3-3) | 0.24 |

### Primary outcomes

#### STN electrophysiology is similar under ketamine and saline

In all cases, we succeeded to identify the electrophysiological characteristics of the STN, including entry and exit from the nucleus, and motor and limbic subdomains, in both anesthesia protocols. Supplementary Figure 1E and F show examples of spiking activity and STN features of two STN trajectories. Figure 2 shows the average STN features of both groups. NRMS and PSD signatures showed similar patterns, while beta oscillation intensity showed a trend towards higher beta power in the ketamine group (Fig. 2A-B); however, this trend did not reach statistical significance. STN length was similar between ketamine and saline groups (0.2 (−0.415, 0.815) mm - *absolute difference* (95% *CI*); Fig. 2C, D.). There was no significant difference in NRMS peak and AUC, percentage of DLOR, DLOR length (Fig. 2C) and power spectrums, including theta, alpha, gamma and HFOs (Fig. 2D).

**Figure 2.**
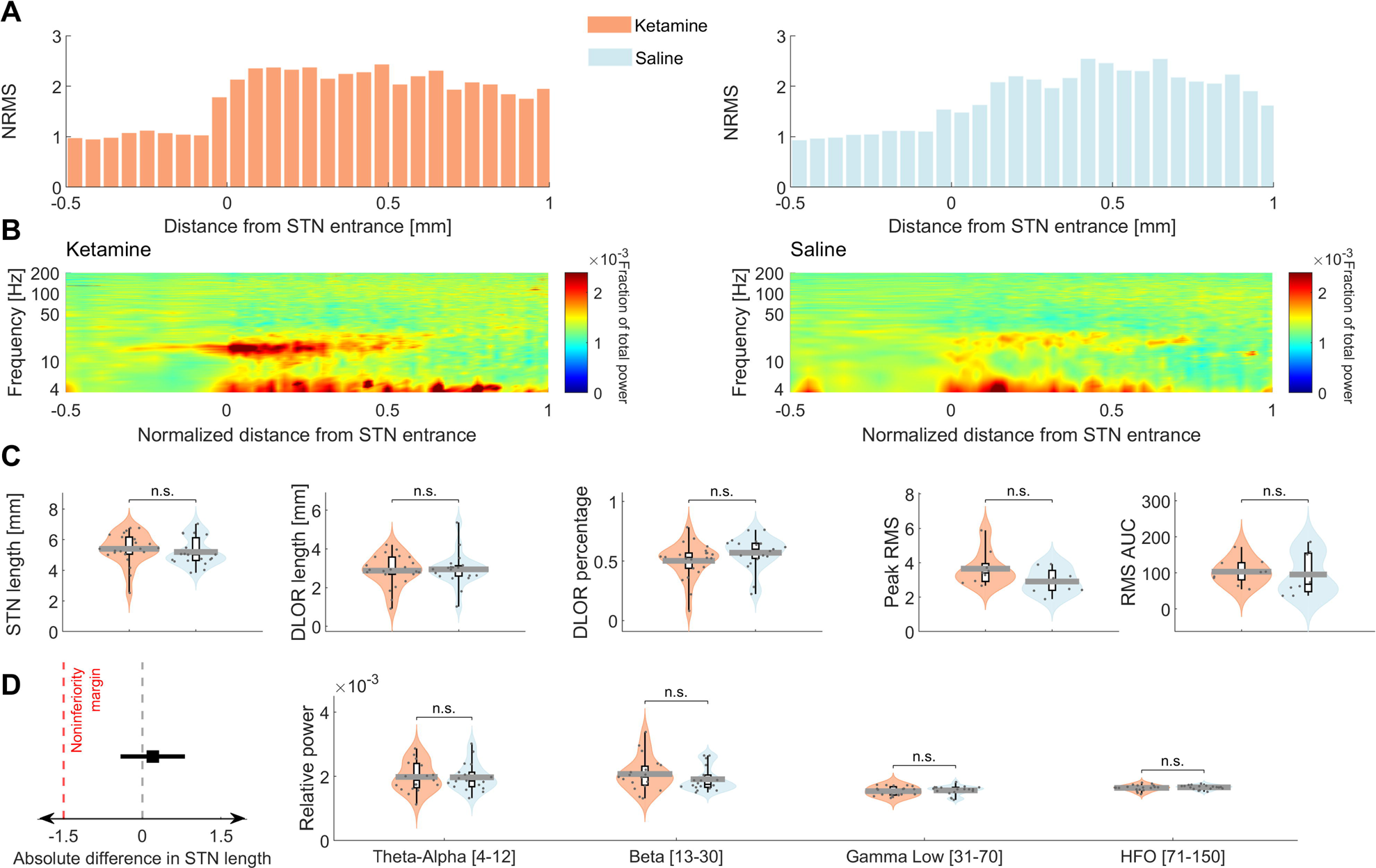
STN electrophysiology under ketamine sedation was noninferior to STN electrophysiology under saline. Ketamine group is indicated in light orange and saline group is indicated in light blue. **(A)** Average normalized root mean square (NRMS) of ketamine (*left*) and saline (*right*) groups along the recorded subthalamic nucleus (STN) trajectory. Data is normalized by the total STN length of each patient. **(B)** Spectrogram of STN trajectory in ketamine (*left*) and saline (*right*) groups. Data is normalized by total power at each depth. X-axis is normalized as in **A**. Color code indicate the fraction of power of frequency bin out of the total power. **(C)** From *left* to *right*: STN length in mm; DLOR length in mm; DLOR percentage out of total STN length; peak NRMS; NRMS area under curve. **(D)** From *left* to *right*: Noninferiority test for STN length between ketamine and saline groups; relative power for 4 frequency bands: 4-12 Hz, 13-30 Hz, 31-70 Hz, 71-150 Hz. Significance assessed with Mann-Whitney test. N.S – p-value >0.05. RMS – root mean squared; DLOR - dorso-lateral oscillatory region.

#### Anesthesia level and patient cooperation during the surgery

The patients’ anesthesia depth and level of consciousness were continuously monitored through clinical observation and the utilization of the Bispectral index, as shown in Fig. 3A. At baseline, all patients exhibited BIS values consistent with an awake stage (ketamine group 92.6 (91.5-96.2) vs. saline group 92.8 (91.4-94.3), *p* = 0.6). During the first and second propofol sedation sections, BIS decreased to the level of light/moderate sedation (79.15 (73-81.72)), and there was no difference between the two groups (*p* = 0.3). During ketamine/saline administration, BIS values increased again to awake level (ketamine group 88.82 (87.31-90.55) vs. saline group 89.14 (88.06-89.95), *p* = 0.8). Patients of both groups were alert enough to fully cooperate during MER and clinical assessment. Some patients from the ketamine group reported a pleasant, dream-like experience during surgery, such as boat trips or odyssey-like journeys. None of the patients reported a negative hallucinatory experience either during or after surgery. Most patients had mild euphoria under ketamine and were relaxed.

**Figure 3.**
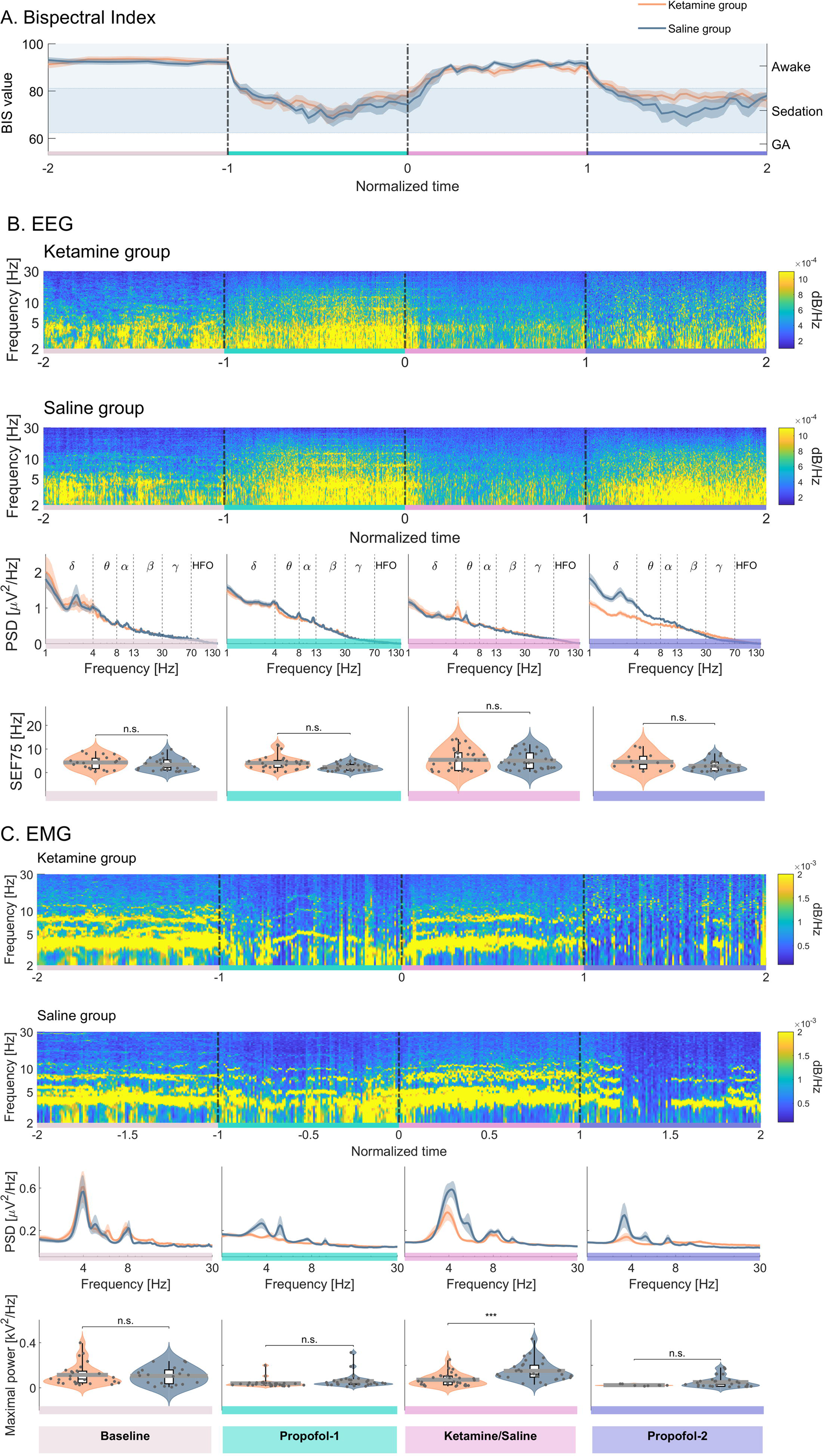
EEG and EMG analysis showed similar patterns in the ketamine and saline groups. Ketamine group is indicated in orange and saline group is indicated in blue. Experimental stages: baseline (beige), propofol-1 (green), ketamine/saline (pink), propofol-2 (purple). **(A)** BIS values during the whole experiment, time normalized. **(B)** EEG analysis. From *top* to *bottom*: spectrograms for ketamine and saline groups; mean power spectral density comparisons for experimental stages, from 1 to 150 Hz - delta, theta, alpha, beta, gamma and high-frequency oscillations; spectral edge frequency 75% comparisons calculated for frequency range 1-150 Hz. **(C)** EMG analysis. From *top* to *bottom*: spectrograms for ketamine and saline groups; mean power spectral density comparisons for experimental stages, shown from 0.1 to 30 Hz; maximal power comparisons from frequency range between 3 and 6 Hz for different stages. P-values are assessed using 2way-ANOVA with Bonferroni correction. N.S – p-value >0.05; *** - p-value < 0.001. PSD – power spectral density; SEF75-spectral edge frequency 75%.

#### Arousal level during the surgery

In addition to BIS, we employed EEG as another measure to analyze patients’ arousal levels. EEG spectrograms, PSD and PSD SEF75 and frequency band’s AUC comparisons are shown in Fig. 3B and Supplementary Fig. 2. There was no difference in SEF75 between the two groups during the study time frame (Fig. 3B, bottom subplot). However, when looking at each frequency band separately, during ketamine/saline administration, beta band was higher in the ketamine group (*p* = 0.006, Supplementary Fig. 2D). This finding is in line with the tendency for higher power of beta oscillations in the STN of the ketamine group patients (Fig. 2D). Notably, after cessation of ketamine/saline phase and subsequent administration of propofol (propofol-2), the EEG bands exhibited a similar pattern to that observed during the ketamine administration. However, in the saline group, there was an increase in delta to alpha frequencies, accompanied by a decrease in gamma and HFO (Supplementary Fig. 2A-C, E, F).

#### Results of clinical assessment are similar between ketamine and saline groups

To monitor the effect of ketamine on tremor amplitude and duration, we used upper limb EMG analysis shown in Fig. 3C. Before starting sedation, the tremor was easily identified and the average EMG spectrogram shows a robust peak at 4-6 Hz. During propofol-1 and propofol-2 infusions there was a decrease in tremor (4-6 Hz) power in both groups. During ketamine administration, tremor peak power was slightly lower compared to the saline group (*p* = 0.002); however, tremor was still clinically evident under ketamine and could be used for TW assessment in both groups. Regarding rigidity under ketamine, this was evaluated clinically, as there is no good enough objective measure for it. No difference was found between the two groups in rigidity, and the impression was that ketamine does not alter rigidity. Side effects were assessed similarly under ketamine and saline. Instances included capsular activation (ketamine: 33%, saline: 30%, *p* = 0.78), gaze palsy (ketamine: 9.5%, saline: 8%, *p* = 0.8), paresthesia (ketamine: 20.8%, saline: 26.9%, *p* = 0.6), and dysarthria (ketamine: 18.2%, saline: 7.7%, *p* = 0.3). Notably, no significant difference in therapeutic effect assessment was observed between ketamine and saline groups (ketamine: 0.5 (0.44-0.75) mA vs. saline: 0.75 (0.5-0.81) mA, *p* = 0.23).

### Secondary outcomes

#### Hemodynamic stability is superior under ketamine

Hemodynamics analysis is shown at Fig. 4 and Supplementary Figs. 3,4. In both groups, baseline BP was hypertensive (average SPB = 157 mmHg, average MAP = 104.3 mmHg) followed by a decrease to normal ranges during the surgery, probably due to the effect of antihypertensive therapy and reduction of anxiety by the propofol treatment. There was no difference between the groups in average BP values during baseline, propofol-1 and ketamine/saline stages. However, at the propofol-2 stage there was a larger decrease in BP in the saline group compared to ketamine (For SBP: 120.23 (113.68-131.64) mmHg vs 110.47 (101.08-120.77) mmHg, *p* = 0.013; for MAP: 78.67 (74.52-85.77) mmHg vs 73.07 (69.39-76.79) mmHg, *p* = 0.05, ketamine vs saline groups, respectively).

**Figure 4.**
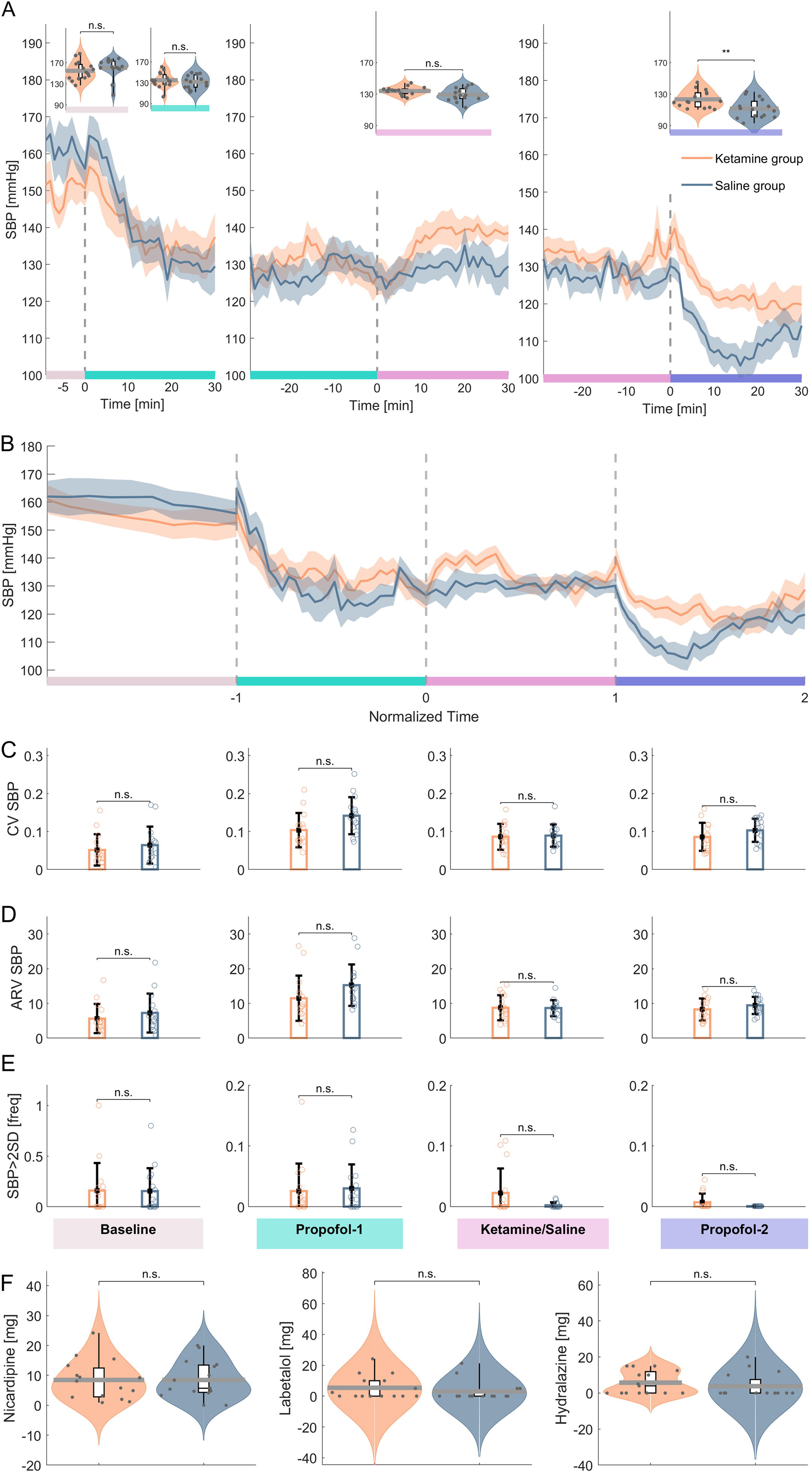
Systolic blood pressure patterns and antihypertensive drug dosage were similar between groups. Ketamine group is indicated in orange and saline group is indicated in blue. Experimental stages: baseline (beige), propofol-1 (green), ketamine/saline (pink), propofol-2 (purple). **(A)** Systolic blood pressure dynamics during the experiment. From left to right: 10-minute baseline measurement – first 30 minutes of propofol administration; last 30 minutes of propofol sedation and first 30 minutes of ketamine/saline administration; last 30 minutes of ketamine/saline and first 30 minutes of propofol-2 sedation. First and last 30 minutes of each stage can overlap. Violin plots on the top – average SBP comparisons between the two groups according to stages. **(B)** Systolic blood pressure during whole experimental period after time normalization. **(C-E)** From *top* to *bottom*: Comparison of CV (coefficient of variation), ARV (Average Real Variability), frequency of hypertensive events (more than 2 standard deviations of SBP) in different experimental stages. **(F)** Comparisons of total dose of antihypertensive drugs for nicardipine, labetalol and hydralazine. Significance assessed with Mann-Whitney test. N.S – p-value >0.05; ** - p-value < 0.01.

Blood pressure variability analysis showed less consistent results. There was no difference in BP variability between the saline and the ketamine groups when separated into different surgical stages (Fig. 4C, D). On the other hand, when taking into account the whole experiment, there was an increase in BP variability for both variability indices in the saline group (For SBP: SV 0.1 (0.09-0.13) vs 0.13 (0.11-0.15), *p* = 0.018; ARV 10.49 (9.05-12.95) vs 11.77 (10.29-16.19), *p* = 0.047; for MAP: SV 0.09 (0.07-0.1), vs 0.12 (0.09-0.14), *p* = 0.025; ARV 5.82 (4.7-7.85) vs 8.47 (5.97-9.2), *p* = 0.028; ketamine vs saline groups, respectively). There was no difference in frequency of hypertensive events between the groups (Fig. 4E). Finally, no difference was found between the two groups in HR analysis (Supplementary Fig.4).

#### Clinical outcomes and surgical accuracy are similar in both groups

In both groups, there was no early cognitive decline after surgery (decrease from preoperative MoCA 0 (−4.71-4.77) %, p = 0.95 for ketamine group; 3.45 (−4.97-10.92) %, *p* =0.22 for saline group). Despite the trend towards more cognitive decline in the saline group, there was no significant difference between the groups (*p* = 0.35; Table 1, Fig. 5A). There was 44% decrease in medication in LEDD after surgery (62.5 (53.6-85.86) % vs 48.53 (36.92-57.31) %, *p* =0.13; ketamine vs. saline, respectively; Fig. 5B). Surgery led to 69% average reduction in UPDRS-III on medication and with stimulation on compared to preoperative off med UPDRS-III (Fig.5C) with no difference in the ketamine/saline groups (*p* = 0.5). There was no difference between the accuracy of microelectrode/lead trajectory (radial error, 0.8 (0.375-1) vs 0.6 (0.325-0.9), *p* = 0.6; ketamine vs saline groups, respectively). No complications were reported in the two study groups, except one patient allocated to the saline group that was subsequently excluded due to peri-lead edema^25^.

**Figure 5.**
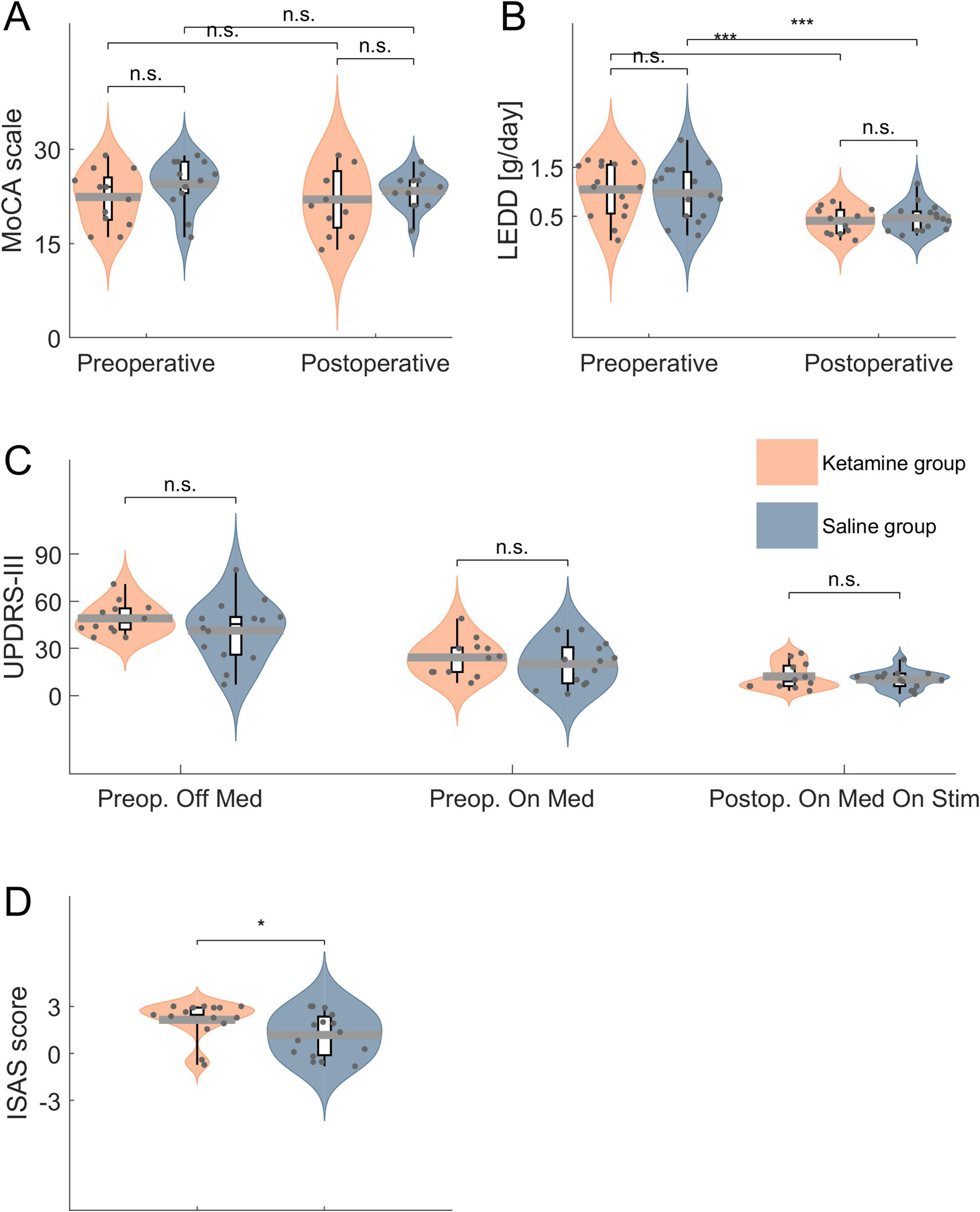
Surgery outcomes were similar, yet patients under propofol-ketamine sedation were more satisfied with their anesthesia. Ketamine group is indicated in orange and saline group is indicated in blue. **(A)** MoCA scales, preoperative and postoperative comparisons. **(B)** LEDD, preoperative and postoperative comparisons. **(C)** UPDRS III, preoperative off-medication, preoperative on-medication and postoperative on-medication and on-stimulation comparisons. **(D)** ISAS score comparisons. **(E)** Modified ISAS score comparisons. MoCA – Montreal Cognitive Assessment scale. LEDD - Levodopa equivalent dose in mg/day. UPDRS III - Unified Parkinson Disease Rating Scale. ISAS – Iowa Satisfaction with anesthesia scale. P-value – Mann-Whitney test, two-tailed for A-D; right-tailed for E, F. N.S – p-value >0.05; * - p-value < 0.05.

#### Patients’ satisfaction is superior under ketamine sedation

When comparing patients’ satisfaction during the “awake” part of surgery, we found that, in line with ketamine’s anxiolytic effects, patients that received ketamine infusion, were more satisfied with their anesthesia (2.45 (2-2.91) vs 1.36 (−0.11-2.34), *p* = 0.032, one-tailed; Fig. 5D). Additionally, we compared every question from the ISAS separately, and found that patients in the ketamine group felt less pain during the surgery (3 (3-3) vs −1 (−2.75-3), *p*=0.013, one-tailed), and less hurt (3 (2-3) vs 2 (−2.75-3), *p* = 0.028, one-tailed). Full ISAS questionnaire with group comparisons can be found in Supplementary Table 1.

## Discussion

This study presents a propofol-ketamine method of moderate sedation for neurosurgical procedures that require active brain state and patient cooperation during clinical assessment, specifically STN DBS for Parkinson’s disease. In this randomized, controlled, double blind study, we show the non-inferiority of ketamine-based conscious dissociative sedation over ‘sham control’ of saline. The study’s findings prove the ability to record basal ganglia multiunit activity without any interference caused by the administered sedative. This approach maintains patients’ calmness and cooperation, leading to excellent lead accuracy and positive patient experience.

### Asleep or not asleep?

The major challenge in DBS surgery is precise lead localization to ensure wide therapeutic window and avoid long term stimulation induced side effects. As many centers move towards “asleep” DBS due to the ease of the procedure for both patient and neurosurgeon, MER and clinical evaluation of the stimulation effects are being abandoned, and anatomical navigation becomes the sole method to assure accuracy. Nevertheless, STN borders and functional subdomains are still not easy to define from standard MR images, and most centers do not use intraoperative-MRI techniques to compensate for brain shift during surgery. Other sedatives used so far in DBS surgery significantly affect MER quality, Parkinson’s disease motor symptoms, and patient cooperation.^2,26–28^ The results of pure anatomic based approaches, or combined anatomic/physiological approaches under hypnotic sedation/anesthesia, are most probably, even if not proven yet, – reduced accuracy.

Our study reveals that low-dose ketamine conscious (dissociative) sedation has multiple advantages. It facilitates intra-operative electrophysiological identification of the STN borders and activity including beta oscillations, it improves patient experience, and increases cooperation by reducing anxiety, discomfort and pain. Moreover, it allows for clinical evaluation during stimulation, as tremor and rigidity are barely influenced by the drug, and therefore results in non-inferior (compared to awake procedure) of lead accuracy and clinical outcome. In addition, ketamine induces fast and stable conscious sedation, with a dissociation-like state, no respiratory depression and optimal hemodynamic stability. In the continuous low dose drip used in this study, it does not cause hallucinations or increase post-surgical complications (cognitive decline, seizures, peri-lead edema, etc.). These results are in agreement with our previous non-human primate and retrospective human studies showing similar STN neurophysiology recordings under ketamine and awake conditions.^14,15^ Overall, it seems to be the most reliable, effective and safe option of all sedatives for DBS.

### Ketamine’s mechanism of action

Ketamine, an *N*-methyl-D-aspartate (NMDA) antagonist, is a dissociative anesthetic agent discovered over 60 years ago, and has been in wide clinical use ever since. Its main characteristics are anesthesia, analgesia, and euphoria. Nowadays it is commonly used in pre-hospital care for pain^29^ and anesthesia of trauma patients,^30^ as well as in treatment of resistant status epilepticus,^31,32^ despite past arguments citing increasing intra-cranial pressure and pro-epileptic effects.^33–36^ More recent studies have even discovered its protective properties in severe traumatic brain injury patients.^37,38^ In recent years, low dose ketamine has also been used for the treatment of major depressive disorder. Moreover, its anxiolytic, cognition enhancing, and anti-epileptic effects have garnered attention in ongoing research.^39–43^

Ketamine acts via NMDA receptors and HCN1 channels to enhance neural activity and produce brain oscillations that are related to its psychotropic effects. Human intracranial recordings revealed that ketamine produces gamma oscillations in the prefrontal cortex and hippocampus, and delta oscillations in the posteromedial cortex. The latter was previously proposed as a mechanism for its dissociative effect.^44^ The potential role of ketamine on depression might be achieved through modulation of AMPA (α-amino-3-hydroxy-5-methyl-4-isoxazolepropionic acid) receptor signaling and function resulting in diminished long term potentiation in the nucleus accumbens of mice lasting at least 7 days.^45^ In our study, we show an increase in low gamma and decrease in delta/theta oscillatory cortical activity in prefrontal EEG recordings during ketamine administration. These results are similar to previously published data.^46,47^ The increase in cortical activity under ketamine is in contrast with robust attenuation of cortical and basal ganglia activity seen under hypnotic sedation (e.g., propofol and dexmedetomidine), even one hour after drug cessation.^28^ In line with the changes in prefrontal EEG activity, most patients under ketamine sedation in our study were in a good mood, sometimes even slightly euphoric, during surgery. This is in agreement with the reported effect of ketamine on mood.^48^ In addition, patients under ketamine did not experience cognitive decline after surgery and did not report amnesia for this part of the surgery. These findings are similar to previous results reported in the literature.^44^

### Hemodynamic effects of ketamine

Ketamine was reported in the past to have unwanted hemodynamic effects for neurosurgery including increase in blood pressure and intra-cranial pressure (ICP). Nevertheless, and in contrast to most neurosurgical procedures, DBS surgery requires average-high blood pressure and ICP to diminish inadvertent effects like pneumocephalus (intracranial air accumulation) and brain shift during the procedure, that affect accuracy of lead localization.

In our study, ketamine resulted in stable blood pressure and heart rate without a decrease in these vital measures, that can be seen with other common sedatives like propofol and dexmedetomidine.^49,50^ The maintenance of hemodynamic stability is of particular importance in DBS surgery, where both increase and decrease in blood pressure may result in suboptimal clinical outcome, including intra-cranial bleeding and reduced lead accuracy,^51^ respectively.

### Dissociative state under ketamine sedation

Another important difference between ketamine and previously used sedatives is the level of consciousness maintained during surgery. Due to the induction of a dissociative state with ketamine, the degree of “brain-wakefulness” of the patient is high (as is reflected from the EEG and high BIS levels). This allows for both clinical testing with a cooperative patient and airway securing without the fear of apnea events. Decreased level of consciousness causes CO_2_ retention and therefore might enhance brain hyperemia and secondary inflammatory injury resulting from the implanted electrodes. These, in turn, might affect postoperative pulmonary complications (atelectasis, aspiration pneumonia, etc.) as well as neurological complications, such as acute delirium, peri-lead edema, and long term cognitive decline.^52,53^ Therefore, the combination of ketamine with interleaving periods of propofol moderate sedation might help to mitigate these unwanted complications and is preferred over a propofol-only regimen.

### Disadvantages of ketamine use

Ketamine itself might also have some unsavory effects, and caution needs to be taken when using it in DBS surgery. Higher ketamine dose might cause undesired hallucinations during surgery, in addition to the dissociation state induced.^54^ This might hamper patient cooperation and result in an overall negative experience for the patient. It is probably a result of the non-selective effect of ketamine on neurotransmitter release in high, sedative dose, and can be avoided by keeping a continuous drip at a low dose.^55^

Another issue is patient variability of response to the drug (using the same dose). Some patients might become sleepy/mildly sedated even when low ketamine dose is used, which, in turn, might affect their cooperation level with stimulation testing.^56^ However, this phenomenon is common with other sedatives as well, and is reversible with dose reduction.

Finally, in some patients ketamine results in a relative decrease in tremor, due to either the decrease in anxiety or non-specific sedative effect. However, in most cases, it is still possible to evaluate tremor during stimulation testing. Nevertheless, this might be of special importance in tremor predominant PD patients and essential tremor patients.

### Study limitations

Our study has a few limitations, mainly the small number of patients enrolled and inability to blind the anesthesiologist and the neurosurgeon. In addition, we used racemic ketamine, while some of the characteristic effects of ketamine were previously described with S-ketamine.^57^ We did not evaluate possible long-term effects of ketamine use on cognition and short- and long-term effects on mood, even though short administration is not supposed to cause long term mood effects.^58^ Also, we did not test differential effect of various doses of ketamine on consciousness level, quality of MER and patients’ subjective experience. Future studies should address all of these questions, as well as the effects of ketamine in DBS for other movement disorders like dystonia and essential tremor, and for other targets, like the globus pallidus and thalamus. Finally, future studies might test the possible use of this regimen in other awake neurosurgical procedures like tumor resection and epilepsy surgery.

## Conclusions

This study strongly supports the use of ketamine-based conscious sedation for awake DBS surgeries. This novel anesthetic regimen offers a safe and unique surgical setup that combines high quality neural recordings with ultimate patient comfort.

## Data Availability

All data produced in the present study are available upon reasonable request to the authors

## Funding

HB was supported by grants of the Silberstein foundation, ISF Breakthrough grant (#1738/22), and the Collaborative Research center TRR295, Germany (#3380/20). IT was supported by a grant of the Israeli Science Foundation (ISF).

## Competing interests

The authors report no competing interests.

## Supplementary material

Supplementary material is available at *Brain* online.

---

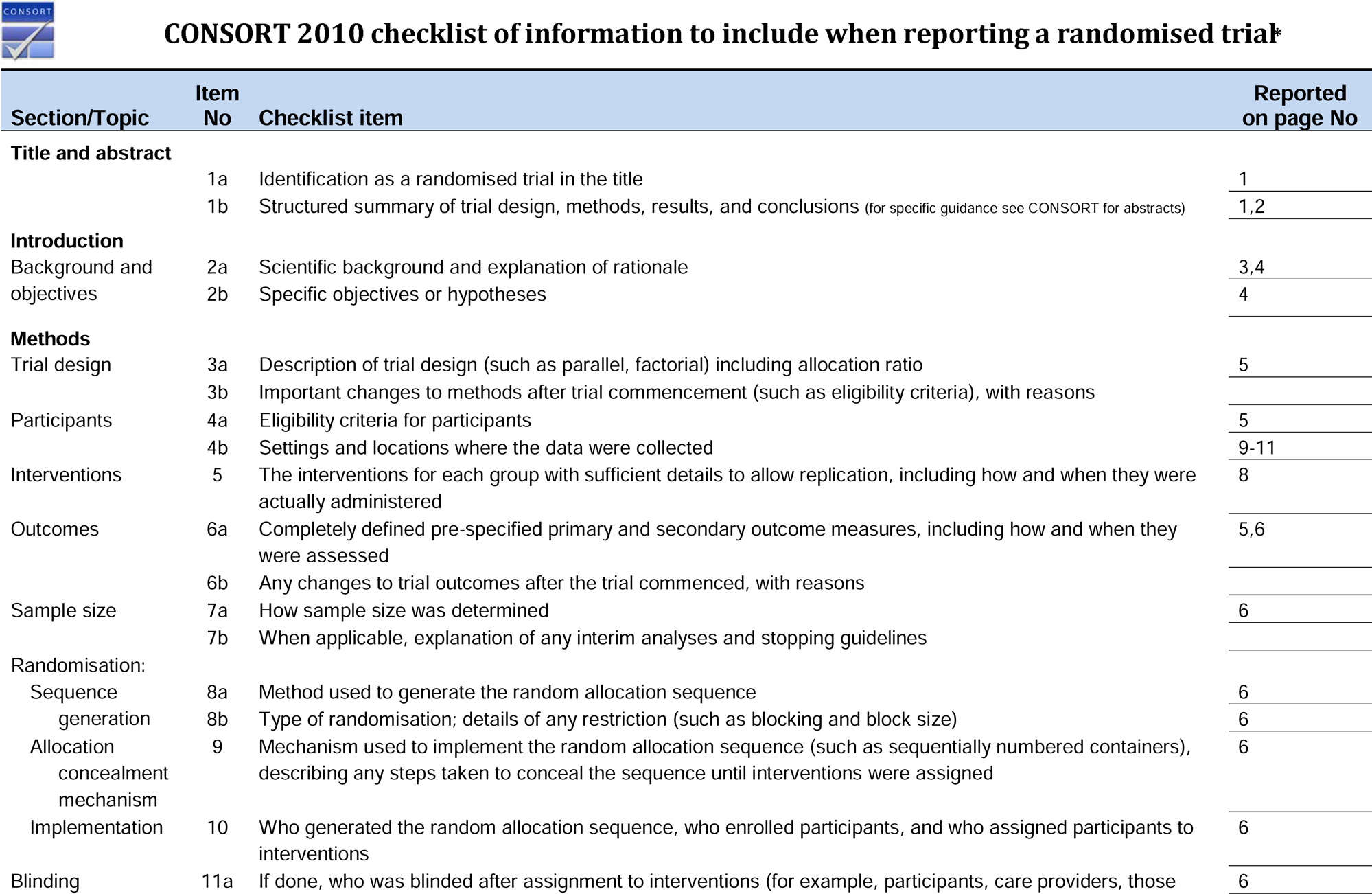
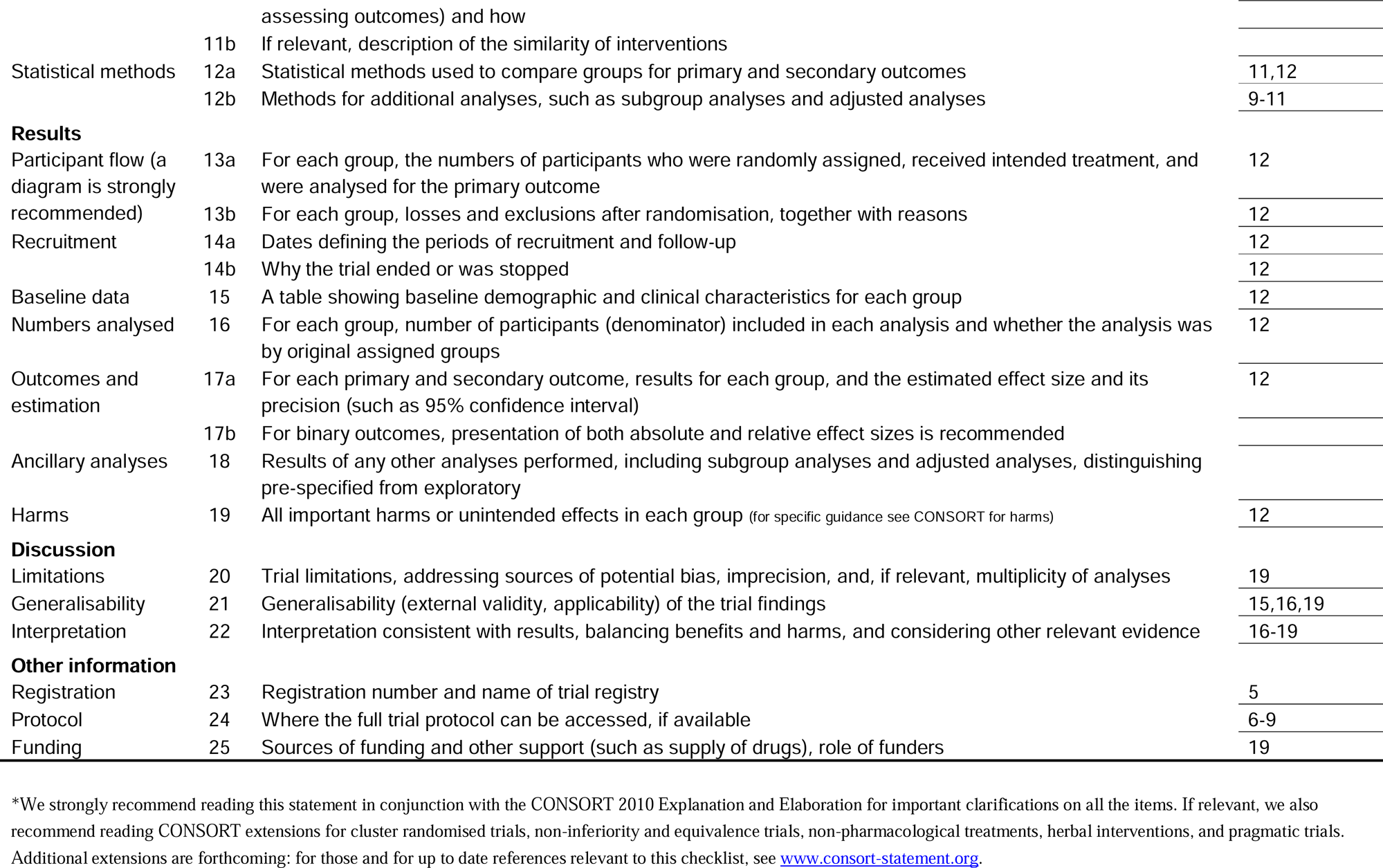

## Notes

### Competing Interest Statement

The authors have declared no competing interest.

### Clinical Trial

NCT04716296

### Funding Statement

This study was funded by Silberstein foundation, ISF Breakthrough grant (#1738/22), the Collaborative Research center TRR295, Germany (#3380/20) and grant of the Israeli Science Foundation (ISF)

### Author Declarations

Ethics committee/IRB 328/50 of Rabin Medical Center, Beilinson Hospital, Israel gave ethical approval for this work

## References

1. Bos MJ, Buhre W, Temel Y, Joosten EAJ, Absalom AR, Janssen MLF. Effect of Anesthesia on Microelectrode Recordings During Deep Brain Stimulation Surgery: A Narrative Review. J Neurosurg Anesthesiol. 2021;33(4):300–307. doi:10.1097/ana.0000000000000673

2. Benady A, Zadik S, Eimerl D, et al. Sedative drugs modulate the neuronal activity in the subthalamic nucleus of parkinsonian patients. Sci Rep. 2020;10(1):14536. doi:10.1038/s41598-020-71358-3

3. Kalenka A, Schwarz A. Anaesthesia and Parkinson’s disease: how to manage with new therapies? Current Opinion in Anesthesiology. 2009;22(3):419. doi:10.1097/ACO.0b013e32832a4b31

4. Martinez-Simon A, Alegre M, Honorato-Cia C, et al. Effect of Dexmedetomidine and Propofol on Basal Ganglia Activity in Parkinson Disease: A Controlled Clinical Trial. Anesthesiology. 2017;126(6):1033–1042. doi:10.1097/ALN.0000000000001620

5. Das S, Matias CM, Ramesh S, et al. Capturing Initial Understanding and Impressions of Surgical Therapy for Parkinson’s Disease. Frontiers in Neurology. 2021;12. Accessed May 14, 2023. https://www.frontiersin.org/articles/10.3389/fneur.2021.605959

6. Vitek JL, Patriat R, Ingham L, Reich MM, Volkmann J, Harel N. Lead location as a determinant of motor benefit in subthalamic nucleus deep brain stimulation for Parkinson’s disease. Front Neurosci. 2022;16:1010253. doi:10.3389/fnins.2022.1010253

7. Shamir RR, Duchin Y, Kim J, et al. Microelectrode Recordings Validate the Clinical Visualization of Subthalamic-Nucleus Based on 7T Magnetic Resonance Imaging and Machine Learning for Deep Brain Stimulation Surgery. Neurosurgery. 2019;84(3):749. doi:10.1093/neuros/nyy212

8. Engelhardt J, Caire F, Damon-Perrière N, et al. A Phase 2 Randomized Trial of Asleep versus Awake Subthalamic Nucleus Deep Brain Stimulation for Parkinson’s Disease. Stereotactic and Functional Neurosurgery. Published online 2021. doi:10.1159/000511424

9. Holewijn RA, Verbaan D, van den Munckhof PM, et al. General Anesthesia vs Local Anesthesia in Microelectrode Recording–Guided Deep-Brain Stimulation for Parkinson Disease: The GALAXY Randomized Clinical Trial. JAMA Neurology. 2021;78(10):1212–1219. doi:10.1001/jamaneurol.2021.2979

10. Rolston JD, Englot DJ, Starr PA, Larson PS. An unexpectedly high rate of revisions and removals in deep brain stimulation surgery: Analysis of multiple databases. Parkinsonism & Related Disorders. 2016;33:72–77. doi:10.1016/j.parkreldis.2016.09.014

11. Khan MF, Mewes K, Gross RE, Škrinjar O. Assessment of Brain Shift Related to Deep Brain Stimulation Surgery. Stereotactic and Functional Neurosurgery. 2007;86(1):44–53. doi:10.1159/000108588

12. Miyagi Y, Shima F, Sasaki T. Brain shift: an error factor during implantation of deep brain stimulation electrodes. Journal of Neurosurgery. 2007;107(5):989–997. doi:10.3171/JNS-07/11/0989

13. van den Munckhof P, Contarino MF, Bour LJ, Speelman JD, de Bie RMA, Schuurman PR. Postoperative Curving and Upward Displacement of Deep Brain Stimulation Electrodes Caused by Brain Shift. Neurosurgery. 2010;67(1):49. doi:10.1227/01.NEU.0000370597.44524.6D

14. Guang J, Baker H, Ben-Yishay Nizri, Orilia Firman S, et al. Toward asleep DBS: cortico-basal ganglia spectral and coherence activity during interleaved propofol/ketamine sedation mimics NREM/REM sleep activity. npj Parkinson’s Disease. 2021;7(1):1–12.

15. Erdman HB, Kornilov E, Kahana E, et al. Asleep DBS under ketamine sedation: Proof of concept. Neurobiology of Disease. 2022;170:105747. doi:10.1016/j.nbd.2022.105747

16. Tamir I, Marmor-Levin O, Eitan R, Bergman H, Israel Z. Posterolateral Trajectories Favor a Longer Motor Domain in Subthalamic Nucleus Deep Brain Stimulation for Parkinson Disease. World Neurosurgery. 2017;106:450–461. doi:10.1016/j.wneu.2017.06.178

17. Tu PH, Liu ZH, Chen CC, et al. Indirect Targeting of Subthalamic Deep Brain Stimulation Guided by Stereotactic Computed Tomography and Microelectrode Recordings in Patients With Parkinson’s Disease. Frontiers in Human Neuroscience. 2018;12. Accessed May 14, 2023. https://www.frontiersin.org/articles/10.3389/fnhum.2018.00470

18. Thompson JA, Oukal S, Bergman H, et al. Semi-automated application for estimating subthalamic nucleus boundaries and optimal target selection for deep brain stimulation implantation surgery. Journal of Neurosurgery. 2018;130(4):1224–1233. doi:10.3171/201712.JNS171964

19. Zaidel A, Spivak A, Shpigelman L, Bergman H, Israel Z. Delimiting subterritories of the human subthalamic nucleus by means of microelectrode recordings and a Hidden Markov Model. Mov Disord. 2009;24(12):1785–1793. doi:10.1002/mds.22674

20. Dexter F, Candiotti KA. Multicenter Assessment of the Iowa Satisfaction with Anesthesia Scale, an Instrument that Measures Patient Satisfaction with Monitored Anesthesia Care. Anesthesia & Analgesia. 2011;113(2):364. doi:10.1213/ANE.0b013e318217f804

21. Valsky D, Marmor-Levin O, Deffains M, et al. Stop! border ahead: Automatic detection of subthalamic exit during deep brain stimulation surgery. Mov Disord. 2017;32(1):70–79. doi:10.1002/mds.26806

22. Appel E, Dudziak R, Palm D, Wnuk A. Sympathoneuronal and sympathoadrenal activation during ketamine anesthesia. Eur J Clin Pharmacol. 1979;16(2):91–95. doi:10.1007/BF00563113

23. Parati G, Stergiou GS, Dolan E, Bilo G. Blood pressure variability: clinical relevance and application. J Clin Hypertens (Greenwich). 2018;20(7):1133–1137. doi:10.1111/jch.13304

24. Mena LJ, Felix VG, Melgarejo JD, Maestre GE. 24LHour Blood Pressure Variability Assessed by Average Real Variability: A Systematic Review and MetaLAnalysis. Journal of the American Heart Association. 6(10):e006895. doi:10.1161/JAHA.117.006895

25. Deogaonkar M, Nazzaro JM, Machado A, Rezai A. Transient, symptomatic, post-operative, non-infectious hypodensity around the deep brain stimulation (DBS) electrode. Journal of Clinical Neuroscience. 2011;18(7):910–915. doi:10.1016/j.jocn.2010.11.020

26. Bos MJ, Alzate Sanchez AM, Smeets AYJM, et al. Effect of Anesthesia on Microelectrode Recordings during Deep Brain Stimulation Surgery in Tourette Syndrome Patients. Stereotactic and Functional Neurosurgery. 2019;97(4):225–231. doi:10.1159/000503691

27. Raz A, Eimerl D, Zaidel A, Bergman H, Israel Z. Propofol Decreases Neuronal Population Spiking Activity in the Subthalamic Nucleus of Parkinsonian Patients. Anesthesia & Analgesia. 2010;111(5):1285–1289. doi:10.1213/ANE.0b013e3181f565f2

28. Amlong C, Rusy D, Sanders RD, Lake W, Raz A. Dexmedetomidine depresses neuronal activity in the subthalamic nucleus during deep brain stimulation electrode implantation surgery. BJA Open. 2022;3:100088. doi:10.1016/j.bjao.2022.100088

29. Sandberg M, Hyldmo PK, Kongstad P, et al. Ketamine for the treatment of prehospital acute pain: a systematic review of benefit and harm. BMJ Open. 2020;10(11):e038134. doi:10.1136/bmjopen-2020-038134

30. Baekgaard JS, Eskesen TG, Sillesen M, Rasmussen LS, Steinmetz J. Ketamine as a Rapid Sequence Induction Agent in the Trauma Population: A Systematic Review. Anesthesia & Analgesia. 2019;128(3):504. doi:10.1213/ANE.0000000000003568

31. Gaspard N, Foreman B, Judd LM, et al. Intravenous ketamine for the treatment of refractory status epilepticus: A retrospective multicenter study. Epilepsia. 2013;54(8):1498–1503. doi:10.1111/epi.12247

32. Fang Y, Wang X. Ketamine for the treatment of refractory status epilepticus. Seizure. 2015;30:14–20. doi:10.1016/j.seizure.2015.05.010

33. Gibbs JM. The effect of intravenous ketamine on cerebrospinal fluid pressure. British Journal of Anaesthesia. Published online 1972. doi:10.1093/bja/44.12.1298

34. Shapiro HM, Wyte SR, Harris AB. Ketamine anaesthesia in patients with intracranial pathology. British Journal of Anaesthesia. Published online 1972. doi:10.1093/bja/44.11.1200

35. Gardner AE, Dannemiller FJ, Dean D. Intracranial cerebrospinal fluid pressure in man during ketamine anesthesia. Anesthesia and analgesia. Published online 1972. doi:10.1213/00000539-197209000-00019

36. Kayama Y, Iwama K. The EEG, evoked potentials, and single-unit activity during ketamine anesthesia in cats. Anesthesiology. Published online 1972. doi:10.1097/00000542-197204000-00004

37. Dengler BA, Karam O, Barthol CA, et al. Ketamine Boluses Are Associated with a Reduction in Intracranial Pressure and an Increase in Cerebral Perfusion Pressure: A Retrospective Observational Study of Patients with Severe Traumatic Brain Injury. Critical Care Research and Practice. 2022;2022:e3834165. doi:10.1155/2022/3834165

38. Zeiler FA, Teitelbaum J, West M, Gillman LM. The Ketamine Effect on ICP in Traumatic Brain Injury. Neurocrit Care. 2014;21(1):163–173. doi:10.1007/s12028-013-9950-y

39. Hudetz JA, Pagel PS. Neuroprotection by Ketamine: A Review of the Experimental and Clinical Evidence. Journal of Cardiothoracic and Vascular Anesthesia. Published online 2010. doi:10.1053/j.jvca.2009.05.008

40. Zarate CA, Niciu MJ. Ketamine for depression: Evidence, challenges and promise. World Psychiatry. Published online 2015. doi:10.1002/wps.20269

41. Taylor JH, Landeros-Weisenberger A, Coughlin C, et al. Ketamine for social anxiety disorder: A randomized, placebo-controlled crossover trial. Neuropsychopharmacology. Published online 2018. doi:10.1038/npp.2017.194

42. Bell JD. In vogue: Ketamine for neuroprotection in acute neurologic injury. Anesthesia and Analgesia. Published online 2017. doi:10.1213/ANE.0000000000001856

43. Pribish A, Wood N, Kalava A. A Review of Nonanesthetic Uses of Ketamine. Anesthesiology Research and Practice. Published online 2020. doi:10.1155/2020/5798285

44. Tian F, Lewis LD, Zhou DW, et al. Characterizing brain dynamics during ketamine-induced dissociation and subsequent interactions with propofol using human intracranial neurophysiology. Nat Commun. 2023;14(1):1748. doi:10.1038/s41467-023-37463-3

45. Yao N, Skiteva O, Zhang X, Svenningsson P, Chergui K. Ketamine and its metabolite (2R,6R)-hydroxynorketamine induce lasting alterations in glutamatergic synaptic plasticity in the mesolimbic circuit. Mol Psychiatry. 2018;23(10):2066–2077. doi:10.1038/mp.2017.239

46. Guang J, Baker H, Ben-Yishay Nizri O, et al. Toward asleep DBS: cortico-basal ganglia spectral and coherence activity during interleaved propofol/ketamine sedation mimics NREM/REM sleep activity. npj Parkinsons Dis. 2021;7(1):1–12. doi:10.1038/s41531-021-00211-9

47. Slovik M, Rosin B, Moshel S, et al. Ketamine induced converged synchronous gamma oscillations in the cortico-basal ganglia network of nonhuman primates. Journal of Neurophysiology. 2017;118(2):917–931. doi:10.1152/jn.00765.2016

48. Kritzer MD, Mischel NA, Young JR, et al. Ketamine for treatment of mood disorders and suicidality: A narrative review of recent progress. Ann Clin Psychiatry. 2022;34(1):33–43. doi:10.12788/acp.0048

49. Forget P, Cata J. Stable anesthesia with alternative to opioids: Are ketamine and magnesium helpful in stabilizing hemodynamics during surgery? A systematic review and meta-analyses of randomized controlled trials. Best Practice & Research Clinical Anaesthesiology. 2017;31(4):523–531. doi:10.1016/j.bpa.2017.07.001

50. Sabertanha A, Shakhsemampour B, Ekrami M, Allahyari E. Comparison of Infusion of Propofol and Ketamine-Propofol Mixture (Ketofol) as Anesthetic Maintenance Agents on Blood Pressure of Patients Undergoing Orthopedic Leg Surgeries. Anesth Pain Med. 2019;9(6):e96998. doi:10.5812/aapm.96998

51. Krauss P, Van Niftrik CHB, Muscas G, Scheffler P, Oertel MF, Stieglitz LH. How to avoid pneumocephalus in deep brain stimulation surgery? Analysis of potential risk factors in a series of 100 consecutive patients. Acta Neurochir. 2021;163(1):177–184. doi:10.1007/s00701-020-04588-z

52. Deng RM, Liu YC, Li JQ, Xu JG, Chen G. The role of carbon dioxide in acute brain injury. Med Gas Res. 2020;10(2):81–84. doi:10.4103/2045-9912.285561

53. Mutch WAC, El-Gabalawy R, Ryner L, et al. Brain BOLD MRI O2 and CO2 stress testing: implications for perioperative neurocognitive disorder following surgery. Critical Care. 2020;24. doi:10.1186/s13054-020-2800-3

54. Powers III AR, Gancsos MG, Finn ES, Morgan PT, Corlett PR. Ketamine-Induced Hallucinations. Psychopathology. 2015;48(6):376–385. doi:10.1159/000438675

55. Bowdle AT, Radant AD, Cowley DS, Kharasch ED, Strassman RJ, Roy-Byrne PP. Psychedelic Effects of Ketamine in Healthy Volunteers : Relationship to Steady-state Plasma Concentrations. Anesthesiology. 1998;88(1):82–88. doi:10.1097/00000542-199801000-00015

56. Zanos P, Moaddel R, Morris PJ, et al. Ketamine and Ketamine Metabolite Pharmacology: Insights into Therapeutic Mechanisms. Pharmacological Reviews. 2018;70(3):621. doi:10.1124/pr.117.015198

57. Molero P, Ramos-Quiroga JA, Martin-Santos R, Calvo-Sánchez E, Gutiérrez-Rojas L, Meana JJ. Antidepressant Efficacy and Tolerability of Ketamine and Esketamine: A Critical Review. CNS Drugs. 2018;32(5):411–420. doi:10.1007/s40263-018-0519-3

58. Lahti AC, Warfel D, Michaelidis T, Weiler MA, Frey K, Tamminga CA. Long-term outcome of patients who receive ketamine during research. Biological Psychiatry. 2001;49(10):869–875. doi:10.1016/S0006-3223(00)01037-4

