## Supplementary material for "Deep brain stimulation surgery under ketamine induced conscious sedation: a double blind randomized controlled trial"

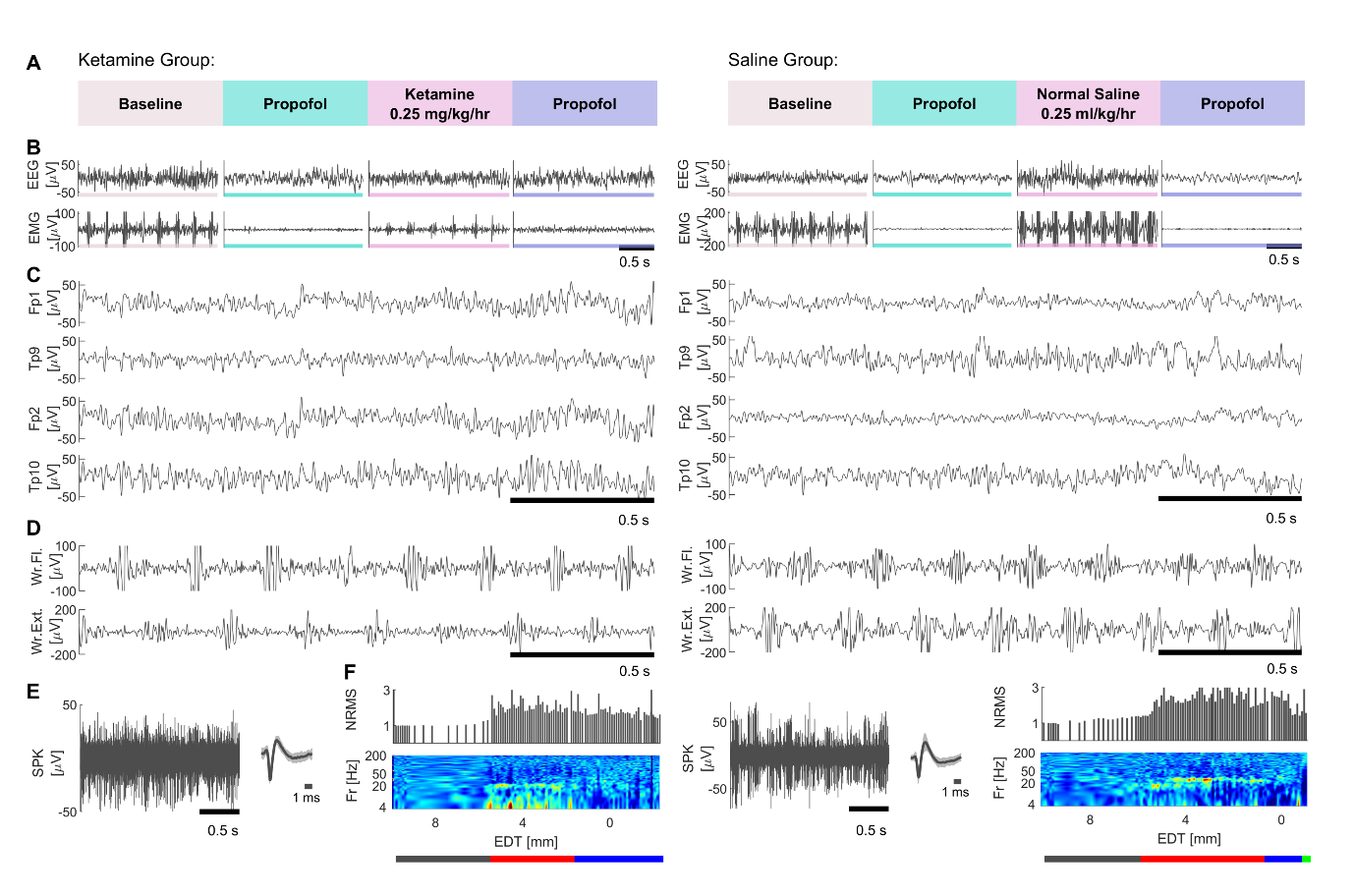

**Supplementary Figure 1 Experimental design and examples of collected data.** Ketamine group traces are shown on the left side, saline group traces – on the right side. Experimental stages: baseline (beige), propofol-1 (green), ketamine/saline (pink), propofol-2 (purple). **(A)** experimental flow for ketamine and saline groups. **(B)** typical examples of EEG (top) and EMG (bottom) traces in the four experimental stages. **(C)** EEG traces during ketamine/saline administration from Fp1, Fp2, Tp9, Tp10 channels. **(D)** EMG traces from wrist flexors (top) and extensors (bottom) during baseline period. **(E)** Spike traces and waveform from microelectrode recordings of STN. **(F)** examples for NRMS, corresponding spectogram from one STN trajectory and Hidden Markov Model results (gray-internal capsule, red – STN dorso-lateral oscillatory region, blue – STN ventro-medial non-oscillatory region, green - Substantia nigra reticulata). STN - subthalamic nucleus, NRMS – normalized root mean squared, EDT – estimated distance to target.

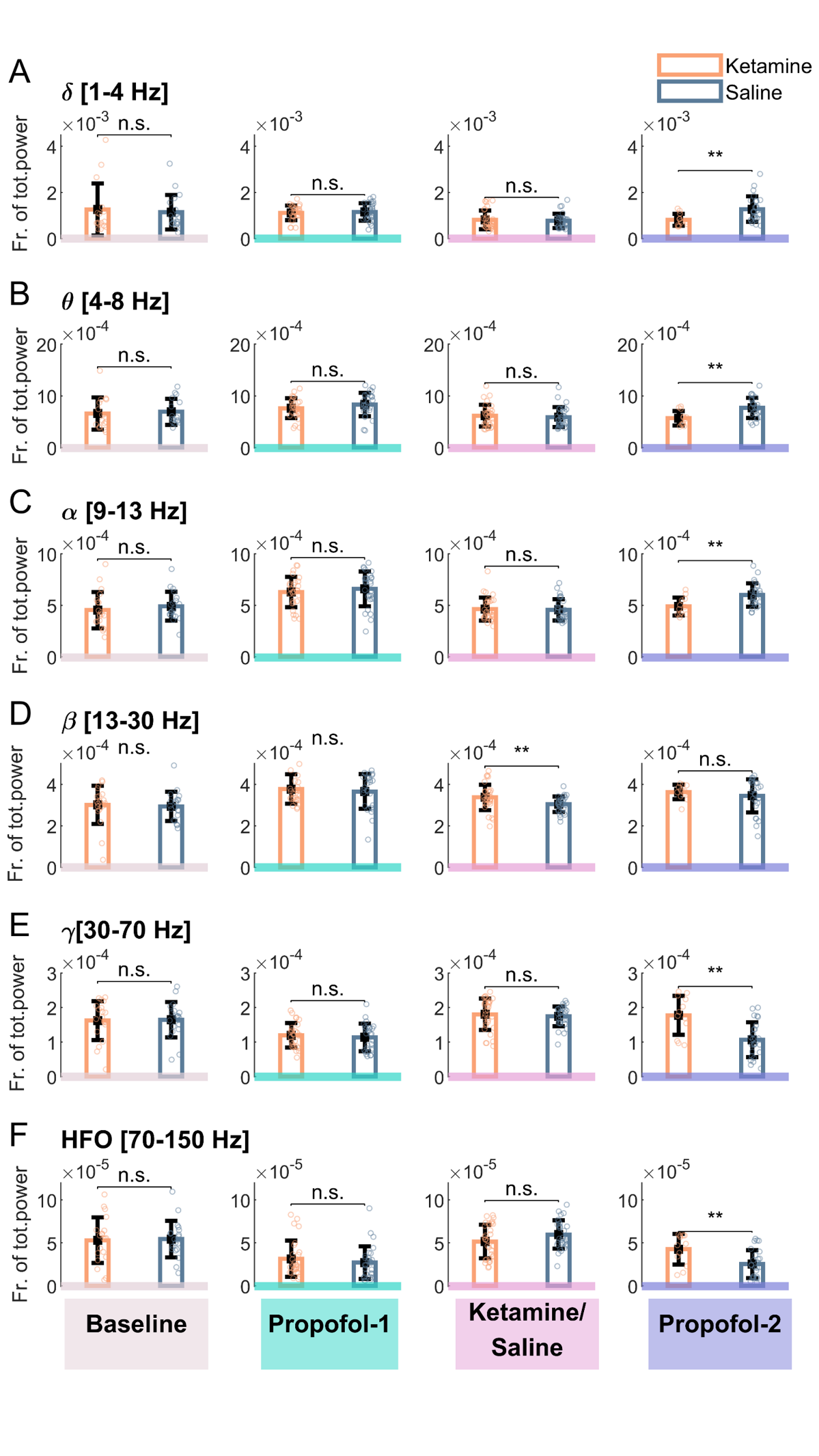

**Supplementary Figure 2 EEG power band comparisons.** Ketamine group is indicated in orange and saline group is indicated in blue. Experimental stages: baseline (beige), propofol-1 (green), ketamine/saline (pink), propofol-2 (purple). Power bands calculated for: **(A)** delta (1-4 Hz); **(B)** theta (4-8 Hz); **(C)** alpha (8-13 Hz); **(D)** beta (13-30 Hz); **(E)** gamma (30-70 Hz); **(F)** high frequency oscillations (70-150 Hz). P-values for comparisons are calculated with Mann-Whitney test (no Bonferroni correction). N.S – p-value >0.05; ** - p-value < 0.01. Y axis labels indicate the fraction of the total power.

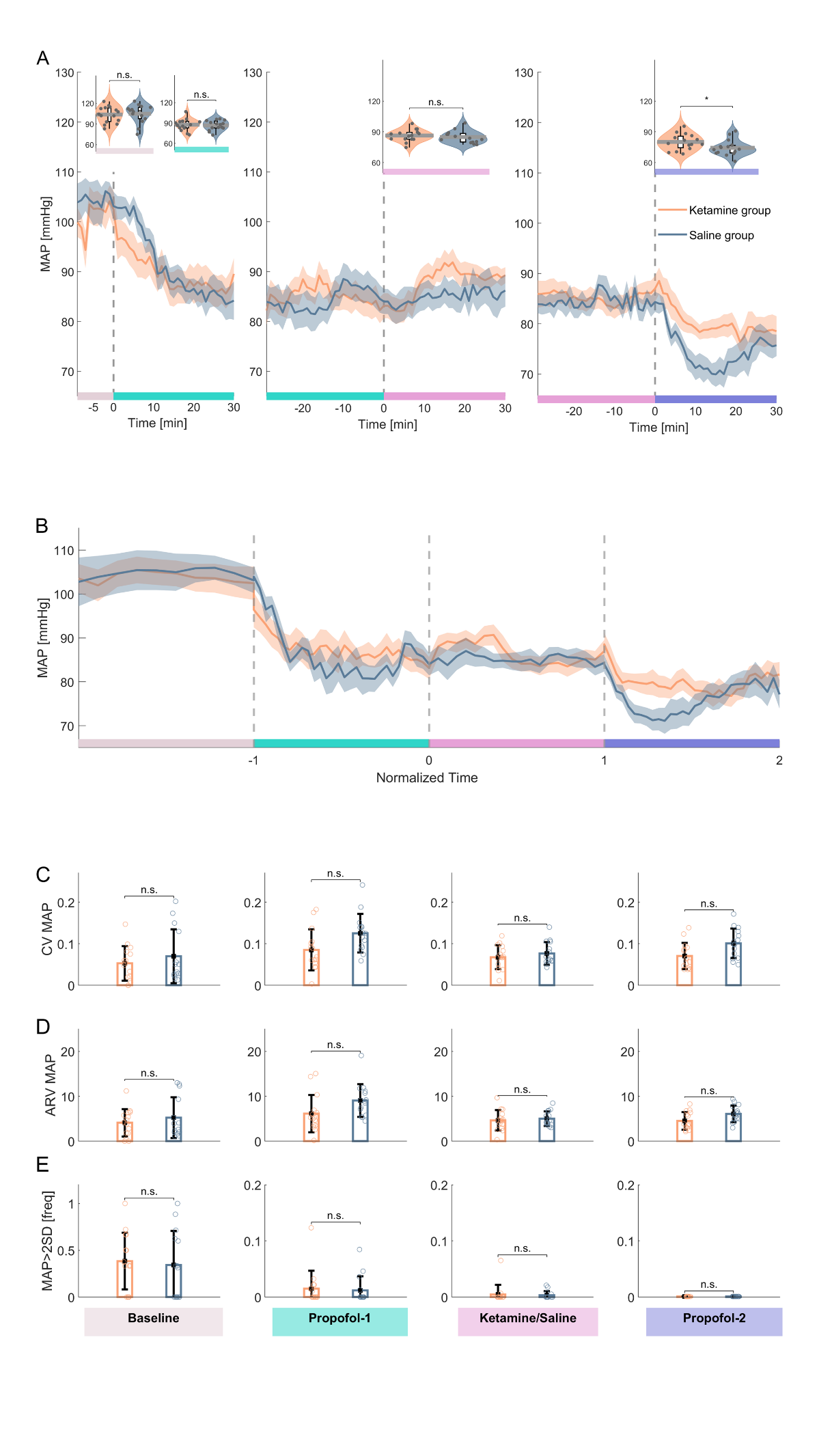

**Supplementary Figure 3 Mean arterial pressure in ketamine and saline groups.** Ketamine group is indicated in orange and saline group is indicated in blue. Experimental stages: baseline (beige), propofol-1 (green), ketamine/saline (pink), propofol-2 (purple). **(A)** Mean arterial pressure (MAP) dynamics during the experiment. From *left* to *righ*t: 10-minute baseline measurement – first 30 minutes of propofol administration; last 30 minutes of propofol sedation and fist 30 minutes of ketamine/saline administration; last 30 minutes of ketamine/saline and first 30 minutes of propofol-2 sedation. Violin plots on the top – average MAP comparisons between the two groups according to stages. **(B)** MAP during whole experimental period after time normalization. **(C-E)** From *top* to *bottom*: Comparison of CV (coefficient of variation), ARV (Average Real Variability), frequency of hypertensive events (more than 2 standard deviation of MAP) in different experimental stages. Significance assessed with Mann-Whitney test. N.S – p-value >0.05; * - p-value < 0.05.

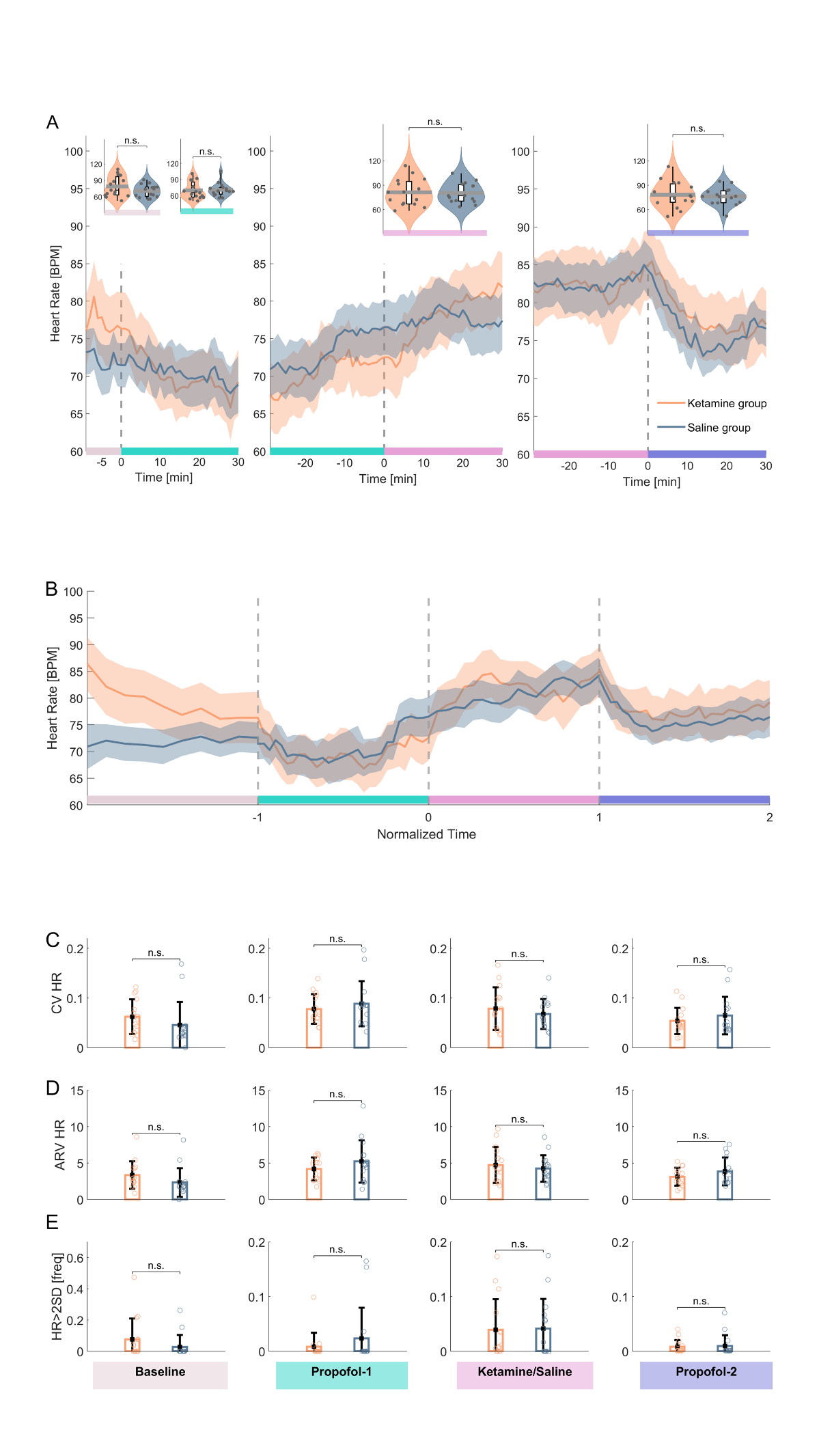

**Supplementary Figure 4 Heart rate dynamics in ketamine and saline groups.** Ketamine group is indicated in orange and saline group is indicated in blue. Experimental stages: baseline (beige), propofol-1 (green), ketamine/saline (pink), propofol-2 (purple). **(A)** Heart rate (HR) dynamics during the experiment. From *left* to *right*: 10-minute baseline measurement – first 30 minutes of propofol administration; last 30 minutes of propofol sedation and first 30 minutes of ketamine/saline administration; last 30 minutes of ketamine/saline and first 30 minutes of propofol-2 sedation. Violin plots on the top – average HR comparisons between the two groups according to stages. **(B)** HR during whole experimental period after time normalization. **(C-E)** From *top* to *bottom*: Comparison of CV (coefficient of variation), ARV (Average Real Variability), frequency of tachycardia events (more than 2 standard deviation of HR) in different experimental stages. Significance assessed with Mann-Whitney test. N.S – p-value >0.05.

**Supplementary Table 1 Anaesthesia satisfaction questionnaire and single question comparisons**

| **Question** | **Ketamine group, N = 15** | **Saline group, N = 15** | **P-value** |
| --- | --- | --- | --- |
| Q1. I threw up or felt like throwing up | 3 (3-3) | 3 (3-3) | 0.458 |
| Q2. I would want to have the same anaesthetic again | 3 (1-3) | -1 (-2.75-3) | 0.052 |
| Q3. I itched | 3 (3-3) | 3 (3-3) | 0.458 |
| Q4. I felt relaxed | 3 (2-3) | 1 (-2.75-3) | 0.052 |
| Q5. I felt pain | 3 (2-3) | -2 (-2.75-3) | 0.045 |
| Q6. I felt safe | | | |
| Q7. I was too cold or hot | 3 (3-3) | 3 (2-3) | 0.28 |
| Q8 I was satisfied with my anaesthetic care | 3 (2.25-3) | 3 (2-3) | 0.155 |
| Q9. I felt pain during surgery | 3 (3-3) | -1 (-2.75-3) | 0.013 |
| Q10. I felt good | 3 (2-3) | 2 (-2-3) | 0.149 |
| Q11. I hurt | 3 (2-3) | 2 (-2.75-3) | 0.028 |
| ISAS total | 2.45 (2-2.91) | 1.36 (-0.11-2.34) | 0.032 |

Data presented as median (IQR). P-value – Mann-Whitney U test (right-tailed), no correction for multiple comparison. ISAS - Iowa Satisfaction Anaesthesia Scale; Q – question.
